# Unraveling temporal dynamics of the post-mortem transcriptome in amyotrophic lateral sclerosis

**DOI:** 10.1101/2025.06.10.25329061

**Authors:** Ting Shen, Barbara E. Spencer, Pavel P. Kuksa, Vivianna M. Van Deerlin, Hemali Phatnani, NYGC ALS Consortium, Edward B. Lee, Corey T. McMillan

## Abstract

**Background:** Human brain transcriptomics contribute an important source of knowledge to our understanding of neurodegeneration but, limited by the cross-sectional nature of these studies captures disease-associated changes at single time points along the disease continuum. Subtype and Stage Inference (SuStaIn) modeling addresses these limitations by inferring temporal gene expression dynamics while also identifying potential transcriptomic subtypes.

**Methods:** We applied a SuStaIn model to improve our understanding of amyotrophic lateral sclerosis (ALS) which has multifaceted heterogeneities. Using bulk RNA-seq data from post-mortem lumbar spinal cord of 172 individuals with ALS and 45 neurologically normal controls, we constructed co-expression modules representing specific molecular pathways, followed by SuStaIn model training to identify molecular subtypes and their transcriptomic trajectories. Subsequently, we extracted subtype-specific hub genes from each subtype, and searched for potential targeted drugs corresponding to these genes. In addition, we validated the model’s reliability and generalizability. Cross-tissue validation applied the model to cervical spinal cord or peripheral blood RNA-seq data.

**Results:** SuStaIn unraveled that more advanced transcriptomic stages were associated with higher microglia and reduced neuron proportions, which mapped onto two ALS subtypes: Immune/Apoptosis/Proteostasis subtype with early immune/apoptotic/proteostatic dysregulation, worse prognosis and higher microglia proportions; Synapse/RNA-Metabolism subtype with early synaptic/RNA-processing deficits, lower male prevalence and more neuron loss. When applying the Lumbar-trained model to cervical spinal cord samples as an independent validation dataset, we observed 71.5% concordance in molecular subtype classification and strong cross-tissue correlation in disease staging (Spearman’s r = 0.76, p < 0.0001). We further identified subtype-specific molecular pathways and hub genes in each ALS subtype. Using the DrugBank database, we retrieved candidate drugs that target these validated hub genes. Low-dose Interleukin-2 (IL-2) therapy may target the Immune/Apoptosis/Proteostasis subtype, and the use of this drug demonstrated a longitudinal reduction in the transcription of genes implicated in the core modules in this subtype.

**Conclusions:** These findings revealed subtype-specific mechanisms underlying ALS heterogeneities, prioritized key genes driving subtyping/staging as potential therapeutic targets, and suggest testable hypotheses that distinct subtypes may exhibit differential responses to targeted interventions. More broadly, we established a framework to decode temporal dynamics from traditionally constrained post-mortem transcriptomic studies.

## Introduction

Human post-mortem tissue studies available through brain donation provide critically important sources of material for understanding the molecular and genomic features of neurodegenerative disease. Particularly, in amyotrophic lateral sclerosis (ALS), multiple molecular mechanisms have been reported to contribute to disease risk and prognosis, some of which include disturbances in RNA metabolism and protein homeostasis, malfunctioning glial cells, axonal disorganization and disrupted transport, mitochondrial dysfunction, endoplasmic reticulum stress, apoptosis and cell survival, as well as exhibiting with a prion-like propagation pattern^1,2^. Specifically, the transcriptomic landscape of brain tissue in ALS has nominated genes associated with many biological pathways including cryptic exon splicing^3^, chromatin accessibility^4^, immune and inflammatory^5^, autophagy and endocytosis^6^, neurological processes such as synaptic transmission and neurotransmitter secretion^5^. Studies also highlight cell-specific features including upregulated genes in microglia and astrocytes and downregulated genes in neurons^7,8^. Moreover, specific autosomal dominant genetic mutations may engage distinct transcriptomic features and molecular pathways, suggesting heterogeneous transcriptomic landscapes across ALS^9,10^. However, a major limitation of these post-mortem studies, that are by definition cross-section and typically end-stage disease, is the lack of clear understanding of the temporal dynamics of transcriptomic features in the ALS human brain. Moreover, the vast majority of transcriptomic evidence focuses on case-control comparisons to identify differentially expressed genes but does not account for potential biological subtypes of ALS.

Recent computational advances have enabled inference strategies from cross-sectional data to address a lack of longitudinal data or identify novel sources of disease heterogeneity but have typically applied either stage-only^11–13^ or subtype-only^14–16^ models. While subtype-only models capture phenotypic heterogeneity, they fail to account for temporal heterogeneity. Conversely, stage-only models describe the dynamic trajectory of disease progression but do not adequately address its heterogeneity. In contrast, the Subtype and Stage Inference (SuStaIn) algorithm has been employed to identify distinct subtypes and extract their progression patterns simultaneously^17^. This model has already been successfully applied to identify data-driven neuropathology-based^18^ and neuroimaging-based^19^ subtypes within ALS-FTD spectrum, allowing for a more nuanced analysis of spatial and temporal disease progression. However, we are unaware of SuStaIn-based strategies for understanding the temporal and heterogenous nature of the human transcriptome.

Here, we sought to leverage bulk RNA sequencing (RNA-seq) data from human post-mortem spinal cord tissue to unravel the temporal dynamics of transcriptomic signatures in ALS, disentangling the disease heterogeneities by transcriptomic subtyping and staging. Our approach is summarized in Fig. 1. We specifically focus on bulk RNA-seq to enable a comprehensive assessment of proportional changes in cell-types across disease stages. With this unsupervised machine learning tool, we were able to identify in lumbar spinal cord tissue two transcriptomic subtypes with distinct temporal transcriptomic trajectories, which reflect the sequential involvement of different molecular pathways during the neurodegenerative process in ALS. The two identified subtypes, related to immune and synaptic functions further generalize well to cervical spinal cord tissue. When applied to blood, the immune subtype is preserved and in a re-analysis of a recent low-dose interleukin-2 (IL-2) trial in ALS we observed longitudinal blood transcriptomic changes in genes implicated in this novel ALS subtype. Together, through novel analytic methods we demonstrate utility in our approach for modeling the post-mortem transcriptome that has translational applications to peripheral blood and clinical trials.

**Fig. 1.**
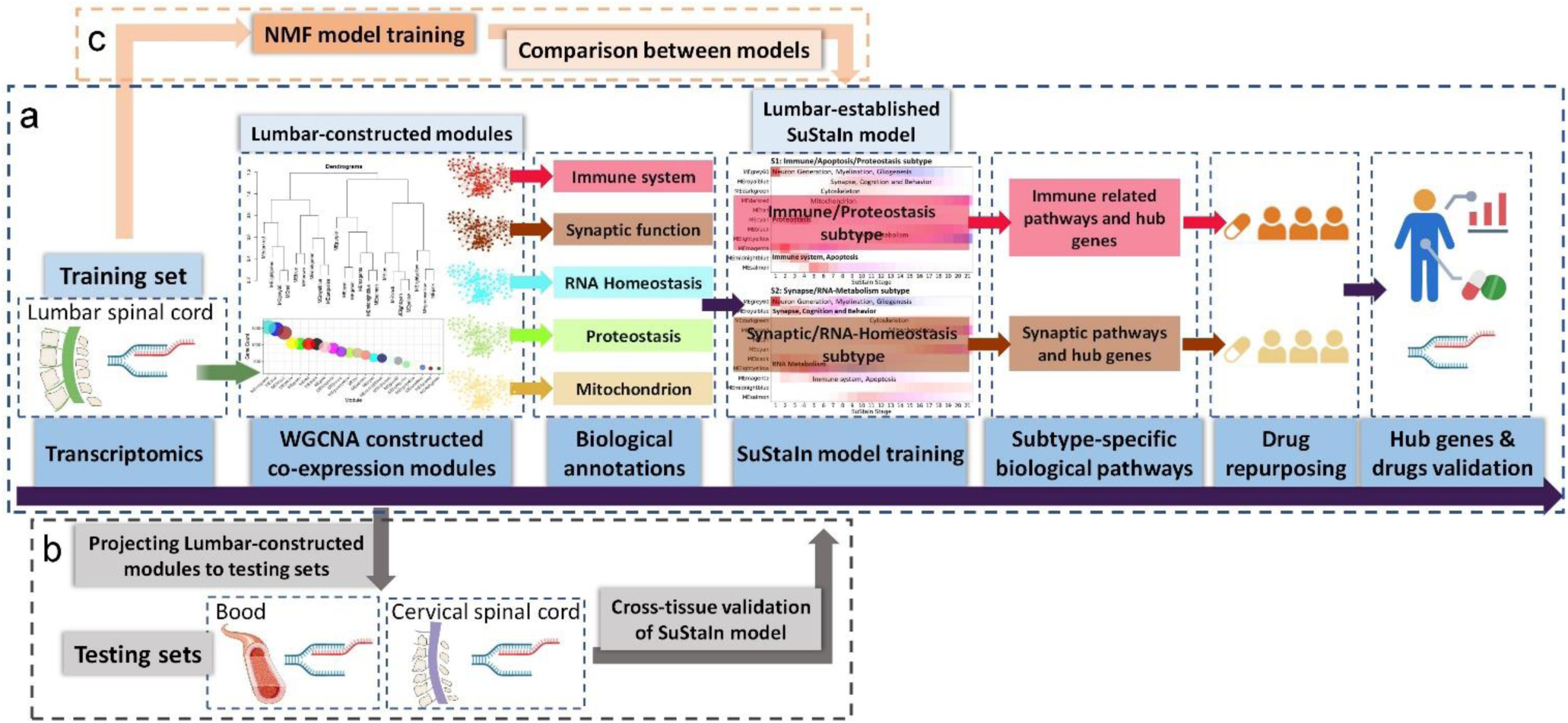
Overview of the study workflow. **a** Construction of the SuStaIn model. A set of co-expression gene modules were constructed using WGCNA based on lumbar spinal cord data (training set). The SuStaIn model was trained, from which subtype-specific hub genes and their related drugs were identified. **b** Cross-tissue validation of the SuStaIn model. Independent testing sets from other tissues were projected onto the modules built from lumbar spinal cord data to validate the generalizability of the SuStaIn model across tissues. **c** Comparison between the SuStaIn model and subtype-only model. The NMF subtype-only model was directly applied to the high-dimensional lumbar spinal cord RNA-seq data and compared with the SuStaIn model. SuStaIn = Subtype and Stage Inference; WGCNA = weighted gene co-expression network analysis; NMF = non-negative matrix factorization.

## Methods

### Study Oversight

The New York Genome Center (NYGC) Amyotrophic Lateral Sclerosis (ALS) Consortium samples presented in this work were acquired through various institutional review board (IRB) protocols from member sites and the Target ALS postmortem tissue core and transferred to the NYGC in accordance with all applicable foreign, domestic, federal, state and local laws and regulations for processing, sequencing and analysis. The Biomedical Research Alliance of New York (BRANY) IRB serves as the central ethics oversight body for the NYGC ALS Consortium. Ethical approval was given and is effective.

### NYGC ALS Consortium cohort

We downloaded post-mortem bulk RNA-seq data of spinal cord from the New York Genome Center (NYGC) ALS Consortium, contributed by eight medical centers. The dataset consists of 366 RNA-seq samples, comprising 216 samples from cervical and 202 from lumbar regions. The dataset included 151 individuals with ALS, 21 with ALS-FTD, and 45 non-neurological controls. A total of 150 individuals have RNA-seq data from both the lumbar and cervical spinal cords, including 113 with ALS, 17 with ALS-FTD, and 20 non-neurological controls. Clinical variables including clinical phenotypes, age at symptom onset, age at death, onset site, sex, and genetic information including the *C9orf72* repeat expansions, as well as technical variables including contributing site, RNA integrity number (RIN), and sequencing platform were extracted from the database (Supplementary Table 1). The disease duration (from symptom onset to death) for ALS/ALS-FTD patients ranged from 6 to 156 months. Individuals with ALS-FTD had shorter disease duration, higher percentage of bulbar onset (50.0%), and higher frequencies of *C9orf72* repeat expansion compared to those with ALS.

### RNA-seq data preprocessing and differential expression analyses

Bulk RNA-seq data processing of NYGC samples are previously described^7,20^. RNA extracted from flash-frozen postmortem tissue was quality-controlled using the Bioanalyzer, and only samples with RIN ≥ 5 were included. Libraries constructed from 500 ng of total RNA were sequenced on Illumina HiSeq 2500 (125-bp paired-end) or NovaSeq (100-bp paired-end) platforms. Gene expression was quantified using the “featureCounts” program (v2.0.2) for GRCh38/hg38 mapped RNA-seq reads^21^. The expression level of each gene was calculated as the sum of raw counts of its exons, where each exon’s total count was derived from the number of non-duplicated paired reads mapped to it. Multi-mapped reads were counted towards expression using weighted/fractional counts (1/x, where x is the number of genomic locations the read has been mapped to). The set of genes quantified for each sample was derived from the GENCODE v37 hg38 annotation using the Genotype Tissue Expression (GTEx) gene model pipeline (https://github.com/broadinstitute/gtex-pipeline). A total of 59,782 genes were quantified for each sample. Infrequently expressed genes were filtered out using a threshold of >= 10 counts in at least 75% of samples^22^. We employed t-distributed stochastic neighbor embedding (t-SNE) to detect potential isolated clusters within the data, potentially influenced by contributing sites and RIN (Supplementary Fig. 1a, b). Normalized counts of remained genes were variance stabilizing transform (VST) transformed using DESeq2 package, and adjusted for contributing site and RIN using Limma package (Supplementary Fig. 1c, d). This procedure was performed in lumbar and cervical spinal cord separately.

Differential expression analyses were then performed by DESeq2. Genes with a false discovery rate (FDR)-corrected p-value < 0.05 were significantly differentially expressed between the individuals with ALS/ALS-FTD and controls (Data file S4).

### Cell-type deconvolution

Cell-type deconvolution of bulk RNA-seq samples was conducted with Scaden, a deep neural network algorithm^23^. Trained on simulated bulk RNA-seq samples derived from snRNA-seq data, Scaden provides robust and precise cell fraction predictions comparable to ground-truth cell composition. We utilized two publicly available snRNA-seq datasets as training sets and established two models. One dataset comprises nuclei from lumbar spinal cord of 9 donors (6 males and 3 females, 35-59 years old), downloaded from the Gene Expression Omnibus (ref1: GSE243077)^24^. Another independent scRNA-seq dataset from spinal cord of 3 donors (ref2: GSE205521) was used for validation. Two different sets of estimated proportions of cell types were generated from two Scaden models separately.

### Weighted gene co-expression network analysis

The preprocessed gene expression data in lumbar spinal cord was used to establish co-expression networks using weighted gene co-expression network analysis (WGCNA) package^25^. A soft thresholding power was selected by pickSoftThreshold function to achieve a scale-free topology fit index > 0.8. We constructed a topology overlap matrix (TOM) for hierarchical clustering analysis. We used a dissimilarity threshold of 0.25 to merge similar modules, and identified 22 modules (Data file S1). The module eigengene (ME), defined as the first principal component of a module, was used to represent overall expression level in this module. Of these, 13 modules showed significantly differential expression in ALS/ALS- FTD compared to controls, with 6 modules being down-regulated and 7 modules up-regulated (Fig. 2a, b).

**Fig. 2.**
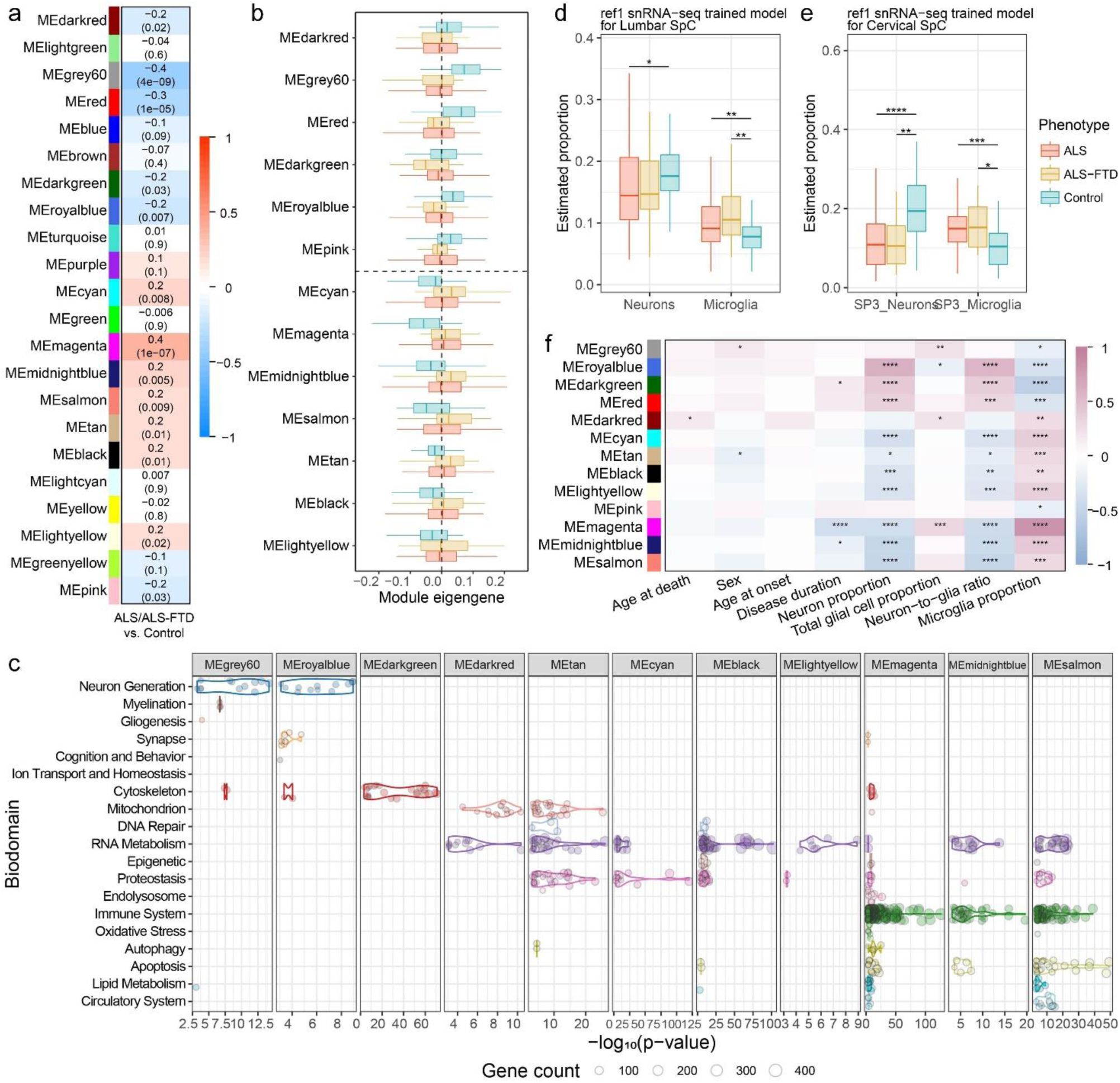
Results of co-expression module construction and cell-type deconvolution. **a** Expression patterns of the 22 modules in ALS/ALS-FTD patients compared to controls. The numbers at the top denote the t-statistics derived from the comparative analysis, whereas the numbers at the bottom signify the *P* values. Blue represents down-regulated and red indicates up-regulated modules in ALS/ALS-FTD relative to controls. **b** Module eigengenes of 13 significantly differentially expressed modules across different clinical phenotypes. These modules exhibit significant expression differences (*P* < 0.05) between ALS/ALS-FTD and controls. **c** Biological annotation of each module. Each dot represents a GO term within a given biodomain. Color of the dots represent different biodomains. The x-axis indicates the significance of term enrichment, and the dot size indicates the number of genes in that GO term and in the module. **d-e** Cell-type proportions estimated by using lumbar snRNA-seq reference (ref1) in bulk RNA-seq sample from SpC across different clinical phenotypes. **f** Correlations of differentially expressed modules with clinical features and estimated cell-type proportions. \**P* < 0.05, \*\**P* < 0.01, \*\*\**P* < 0.001. SpC = spinal cord.

Module membership (MM) was calculated as the correlation between an individual gene and its module’s ME. Genes with high MM are strong representatives of overall module expression. Gene significance (GS) represents the correlation between an individual and group features. To identify key genes within modules that may drive subtyping and staging, we calculated the GS using features such as subtype assignment and stage. Hub genes within modules were identified using a screening criterion of GS > 0.2 and MM > 0.8. We then applied the top 1,000 genes enriched in each cell type as cell-type markers^26^, and identified overlapped genes between these markers and hub genes within each module. For each module, we generated 10,000 random gene sets matching its hub gene counts from the whole gene pool used for WGCNA analysis, and counted overlaps with each cell-type marker set to obtain a null distribution of overlapping gene numbers. The p-value for cell-type enrichment was derived by comparing the actual overlap against this distribution, with significance indicating that module hub genes are enriched for the corresponding cell-type signature^27^.

We used the WGCNA “modulePreservation” function to evaluate whether the Lumbar-constructed modules are preserved in cervical spinal cord dataset. Permutation testing (1,000 times) generated Zsummary statistics, where Zsummary > 10 indicates strong preservation, 2 < Zsummary < 10 indicates moderate preservation, and Zsummary < 2 were non-preserved^28^. All 22 modules showed Zsummary > 10, then we projected the Lumbar-constructed modules onto the cervical dataset and calculated cervical MEs.

### Functional enrichment analyses

To test whether each module map to specific biological pathways, functional enrichment analyses were conducted using gprofiler2^29^. Significant enrichment in Gene Ontology Biological Process (GO:BP) terms was determined using the g:SCS (Set Counts and Sizes) method for multiple testing correction^30^, with a significance threshold of *P*<0.05. Biological annotation of modules centered on biodomains that characterize the ALS/FTD endophenotypes^31,32^. We adopted a published strategy^33^ to filter out GO terms that were too general to be mapped into specific endophenotypic space, or mapped to high-order positions within the ontology, retaining GO terms categorizable into relevant biodomains. The significant GO terms identified for each module were assigned to specific biodomains, thus each module represented specific biological pathways, as the top significant terms and majority of the terms predominantly associated with specific biodomains.

### SuStaIn modelling and validation

The MEs of modules were z-scored, adjusting for age at death and sex, with non-neurologic controls as reference group. For modules exhibiting down-regulation, the z-scores were inverted (multiplied by -1) to ensure all biomarkers are indicative of up-regulation in disease. These z-scored MEs served as input data for SuStaIn model (Supplementary Fig. 2a), which combines clustering and disease progression modelling to identify subtypes with distinct transcriptomic trajectories. Using a 10-fold cross-validation, we selected the optimal number of subtypes based on the out-of-sample log-likelihood and cross-validation information criterion to better balance the model complexity with accuracy (Supplementary Fig. 2b). Transcriptomic progression of each subtype was modeled as a sequence of z-score events using an event-based model, in which each module follows a piecewise linear trajectory over a common timeframe^17^, with each event corresponding to a module transitioning from normal to abnormal expression levels. For the inclusion of z-score events, we utilized z-score waypoints of 1, 2, and 3 for each module, while excluding those z-score events where less than 10 subjects surpassed that z-score waypoint. The final point of this progression trajectory is marked by the maximum z-score, designated as 2, 3, or 5 depending on the maximum z-score event being 1, 2 or 3, respectively^17^. The number of stages is determined by the number of biomarkers and z-score events, which is calculated as the sum of all final z-score events across all biomarkers. Each subtype exhibited distinct transcriptomic progression patterns, initiating with alterations in specific molecular pathways followed by sequential involvement of other pathways in subtype-specific sequences. Each stage represented a module reaching a new expression level, reflecting dynamics of specific pathways. For each subject, the SuStaIn model calculated a probability value to each subtype and stage (Supplementary Fig. 2c, d). The assignment strength was determined by their maximum probability of belonging to one of the subtypes, thereby enabling their assignment to a specific subtype and stage within trajectory of this subtype.

### Lumbar-trained SuStaIn model validation across multiple tissues

Cervical spinal cord RNA-seq data from the same cohort served as an external validation set for the SuStaIn model. We projected the highly preserved Lumbar-constructed modules onto cervical dataset, and calculated the cervical MEs. The Lumbar-trained SuStaIn model fitted to the cervical spinal cord dataset was used to infer transcriptomic subtype and stage in cervical spinal cord. Subtype stability was determined as the proportion of individuals who were assigned to the same subtype.

We further validated this model using peripheral blood transcriptomic data from an independent cohort comprising 397 ALS and 645 controls^34^. Blood RNA-seq dataset was processed with consistent normalization and quality control procedures. We evaluated the preservation of Lumbar-constructed modules, projected these modules onto the blood transcriptomic data, and calculated the blood MEs. The Lumbar-trained SuStaIn model was directly applied to classify blood samples into molecular subtypes. Permutation testing (3,000 iterations) was performed by randomly shuffling the MEs across samples, then applying the SuStaIn model to the permuted data to generate null distributions of subtype proportions. Statistical significance was assessed by comparing the observed subtype frequencies against these null distributions, evaluating whether the observed classifications were likely to occur by chance.

### Non-negative matrix factorization clustering

We further conducted comparative analysis using non-negative matrix factorization (NMF) clustering as an unsupervised subtype-only model^35,36^, on the lumbar spinal cord dataset. Using the NMF package^37^ with non-smooth NMF (nsNMF) method at a factorization rank of 2 and 200 iterations, we conducted 11 rounds of NMF clustering to ensure robust subtype assignment. For each individual, the NMF subtype with a simple majority was selected. The basis matrix, representing gene contributions to each component, was averaged across the 11 clustering replicates. Representative genes for each subtype were selected by ranking genes based on their contributions to the subtype. We selected the top 2,000 genes for each subtype and identified unique and common genes of the NMF-derived subtypes. Subsequently, we conducted functional enrichment analysis on the unique and common genes to explore the biological annotations associated with the two NMF-derived subtypes.

### Identification of potential therapeutic strategies

To translate findings into potential therapeutic insights, we systematically screened the Coremine Medical database to identify potential drugs targeting identified genes^38^. This platform computationally mines millions of biomedical publications and databases to extract and score relationships between biomedical concepts. The top hub genes from core modules (magenta, cyan, royalblue, black) were queried against the database to retrieve pharmacological compounds with high association scores. The results were visualized using Cytoscape software. Furthermore, to assess potential drugs targeting the hub genes, we queried the DrugBank database for drugs known to interact with their corresponding protein products^39^.

Moreover, to investigate transcriptional response to potential therapeutic agents, we reanalyzed transcriptomic data from a clinical trial where ALS patients received subcutaneous low-dose IL-2 injections once daily for 5 days every 28 days, totaling three treatment cycles^40^. Using microarray gene expression data from peripheral leukocytes collected at baseline (Day 1) and post-first cycle (Day 8), we evaluated three WGCNA-constructed modules (magenta, midnightblue, salmon) for their response to IL-2 treatment.

### Statistical analysis

The statistical analysis was conducted with R statistical software version 4.2.0. Normality of variable distribution was tested using Shapiro-Wilk normality test. Continuous variables with normal distribution were compared using two-sample t-test, while Mann-Whitney test was utilized for comparing variables with non-normal distribution. For comparison of categorical variables, chi-squared test or Fisher exact test was employed. After identifying subtypes, we compared clinical and genetic features, cell-type proportions, MEs of modules across subtypes. Correlation analyses were conducted between SuStaIn stages and clinical features, cell-type proportions, MEs of modules. All statistical analyses will be considered significant at a threshold of *P*<0.05.

## Results

### RNA-seq data processing, module construction, and cell-type deconvolution

We report an initial analysis of bulk RNA-seq of lumbar spinal cord from 151 ALS, 21 ALS-FTD and 45 neurologically normal controls (Supplementary Table 1), which was adjusted for RIN and contributing sites (Supplementary Fig. 1). We then used WGCNA to derive 22 transcriptomic modules (Fig. 2a, Supplementary Fig. 3a-d, and Data file S1). Of these, 13 modules showed significantly differential expression in ALS/ALS-FTD compared to controls, with 6 modules being down-regulated and 7 modules up-regulated (Fig. 2a, b). Functional enrichment analyses were conducted for each module (Data file S2), then we mapped 11 out of these 13 significant modules to identifiable biodomains (Fig. 2c, and Supplementary Fig. 3e). The grey60 and royalblue modules were enriched in “Neuron Generation” biodomain. The grey60 module was linked to “Myelination” and “Gliogenesis”, while royalblue module was also associated with “Synapse” biodomain. The darkgreen module related to “Cytoskeleton”, and darkred module was more enriched in “Mitochondrion” biodomain. The cyan module was involved in “Proteostasis”, while black and lightyellow modules were linked to “RNA Metabolism”. The tan module related to “Mitochondrion”, “Proteostasis” and “RNA Metabolism”. The magenta, midnightblue and salmon modules were primarily enriched in “Immune System”, with the salmon module also involved in “Apoptosis”.

Bulk RNA-seq data were deconvolved using Scaden to estimate cell-type proportions ^23^, serving as indicators of neurodegeneration and neuroinflammation. Microglia proportions were higher, while neuron proportions were lower in ALS/ALS-FTD than controls in both lumbar and cervical spinal cord (Fig. 2d, e, and Supplementary Fig. 4a). Estimated cell-type proportions were highly correlated between two independent snRNA-seq references (Supplementary Fig. 4b-d), thus we mainly reported the results of lumbar reference. As shown in Fig. 2f, the grey60 module positively correlated with glial cell proportion (r=0.23, *P*=0.006), whereas the Synapse-related royalblue module exhibited a strong inverse correlation between its down-regulated expression and neuron proportion (r=0.60, *P*<0.0001). The magenta, midnightblue, and salmon modules, enriched in immune-related pathways, correlated with microglia proportions. Notably, the magenta module additionally exhibited negative correlation with disease duration (r=-0.39, *P*<0.0001).

### Novel transcriptomic subtypes of ALS lumbar spinal cord

SuStaIn algorithm was applied to the expression files of the 11 empirically-derived modules, identifying two transcriptomic subtypes with distinct transcriptomic dynamic trajectories (Fig. 3a). The z-score ranging from 1 to 3, indicating the degree of gene expression level from mild to moderate to severe. The most noticeable differences between subtypes were the order in which different modules were involved throughout the progression.

**Fig. 3.**
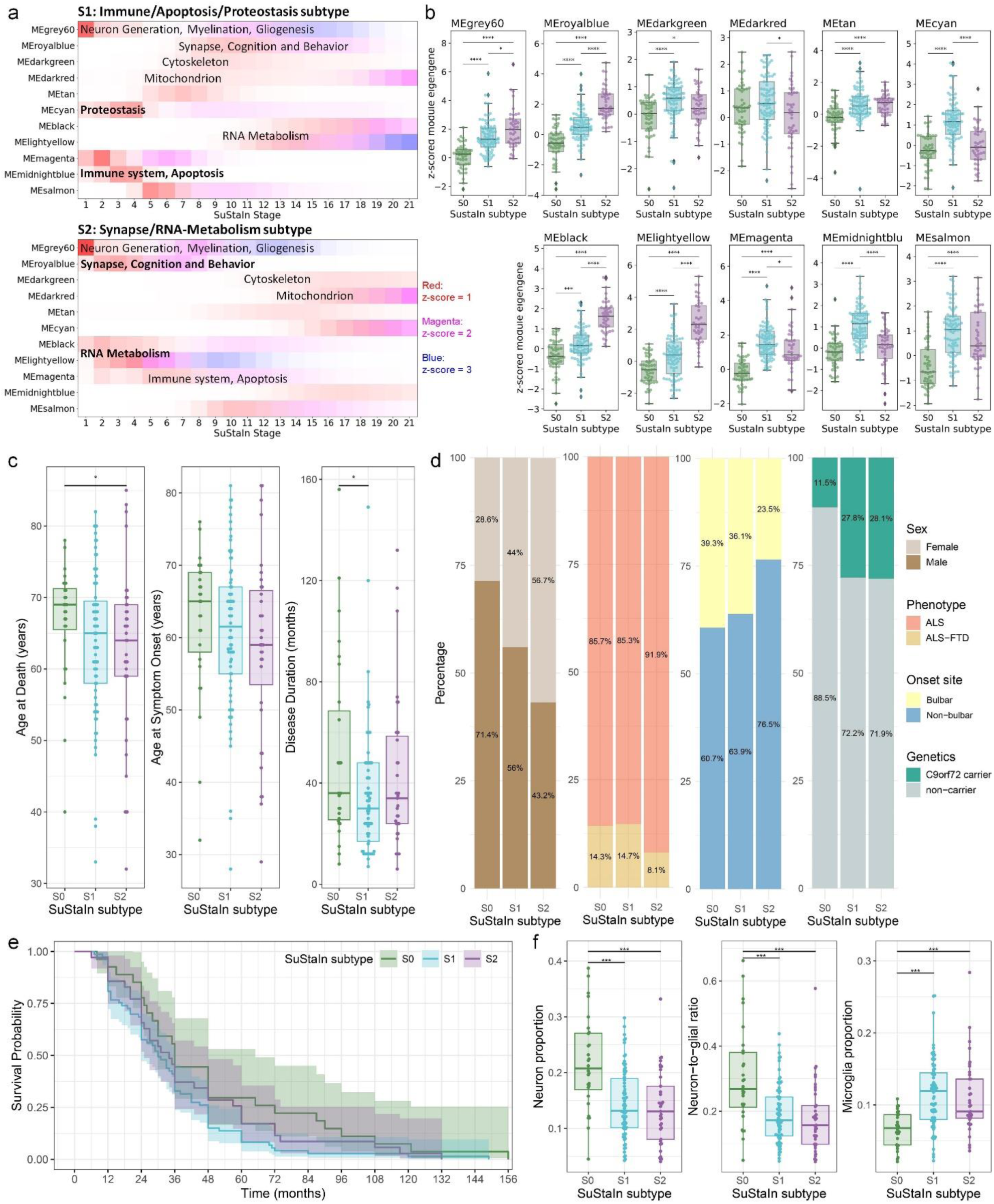
The two transcriptomic subtypes identified by SuStaIn algorithm. **a** Progression trajectories of modules across subtypes. Z-scores represent the dynamic changes of each module along SuStaIn stages. Color gradient indicates the probability of abnormality severity: red (mild), magenta (moderate), blue (severe). **b** Comparisons of z-scored module eigengenes across SuStaIn subtypes for each module. The z-scores of down-regulated modules were multiplied by -1 to ensure that all biomarkers used as inputs for the SuStaIn model are elevated in disease. Each module is labeled with its predominant and specific bidomain characteristics. **c** Bar chart illustrating the distinct distribution of clinical and genetic features across subtypes. **d** Transcriptomic subtypes show significantly younger age and shorter disease duration. **e** Survival analysis across different subtypes. **f** Cellular composition differences across subtypes. Cell-type proportions shown in this figure were estimated by using lumbar snRNA-seq reference (ref1). \**P* < 0.05, \*\**P* < 0.01, \*\*\**P* < 0.001, \*\*\*\**P* < 0.0001. SuStaIn = Subtype and Stage Inference; S0 = Normal-appearing group; S1 = Immune/Apoptosis/Proteostasis subtype; S2 = Synapse/RNA-Metabolism subtype.

The first subtype, we labelled “Immune/Apoptosis/Proteostasis subtype”, initially had early evidence of altered gene expression in “Immune System”, “Apoptosis”, “Proteostasis”, and “Neuron Generation, Myelination, Gliogenesis” biodomains. This subtype subsequently showed abnormality in “Mitochondrion” biodomain at SuStaIn stage 5, followed by involvement of “Cytoskeleton” biodomain at SuStaIn stage 6, “Neuron Generation, Synapse” biodomain at SuStaIn stage 7, and “RNA Metabolism” biodomain at SuStaIn stage 9-11. The second subtype that we labelled “Synapse/RNA-Metabolism subtype” initially involved biodomains of “Synapse”, “RNA Metabolism”, and “Neuron Generation, Myelination, Gliogenesis” at SuStaIn stage 1. This subtype later showed abnormalities in “Immune System” biodomain, progressing at a slower rate compared to the Immune/Apoptosis/Proteostasis subtype. It subsequently showed involvement of “Apoptosis” biodomain at SuStaIn stage 7, the “Cytoskeleton” biodomain at SuStaIn stage 11, and “Mitochondrion” and “Proteostasis” biodomains at SuStaIn stage 13. In addition to these two subtypes with gene expression changes, individuals assigned to SuStaIn stage 0 were labeled as “normal-appearing group”, which showed no detectable transcriptomic changes in these modules. The predominant modules for each transcriptomic subtype exhibited more pronounced abnormal expression levels relative to the other transcriptomic subtype and normal-appearing group (Fig. 3b).

### Characteristics of subtypes

Of the 140 individuals with ALS/ALS-FTD, 75 (53.6%) were categorized as Immune/Apoptosis/Proteostasis subtype, 37 (26.4%) were Synapse/RNA-Metabolism subtype, and 28 (20.0%) were normal-appearing group. The detailed characteristics of subtypes are summarized in Table 1 and Fig. 3c, d. Relative to normal-appearing group, the Immune/Apoptosis/Proteostasis subtype had a shorter disease duration, indicative of worse prognosis, while the Synapse/RNA-Metabolism subtype exhibited an earlier age at death and lower proportion of males. Both subtypes showed a trend toward a higher frequency of repeat expansions in *C9orf72* compared to the normal-appearing group. However, while the Synapse/RNA-Metabolism subtype exhibited a trend toward a lower frequency of bulbar-onset compared to Immune/Apoptosis/Proteostasis, these two subtype groups did not statistically differ from each other on the evaluated clinical factors. Survival analysis also indicated that the Immune/Apoptosis/Proteostasis subtype had a shorter survival time after disease onset (Fig. 3e). The two subtypes showed lower neuron proportions and higher microglia proportions than normal-appearing group (Fig. 3f). While not significant, the Immune/Apoptosis/Proteostasis subtype exhibited higher trends in microglia and neuron proportions compared to Synapse/RNA-Metabolism subtype, consistent with their assignment to the involved biodomains.

**Table 1.** Comparison of clinical characteristics between SuStaIn subtypes.

|  | **S0:**<br>Normal-appearing<br>group | **S1:**<br>Immune/Apoptosis/<br>Proteostasis<br>subtype | **S2:**<br>Synapse/RNA-Metabolism<br>subtype | **Missing**<br>data | **pS0vsS1** | **pS0vsS2** | **pS1vsS2** |
| --- | --- | --- | --- | --- | --- | --- | --- |
| **Subject, n** | 28 | 75 | 37 | - | - | - | - |
| **Age at Death, years** | 66.6 (8.1) | 63.8 (10.1) | 62.0 (12.3) | 0 (0.0%) | 0.10 | **0.03** | 0.44 |
| **Age at symptom onset, years** | 62.3 (10.1) | 60.8 (10.5) | 58.7 (12.6) | 4 (%) | 0.30 | 0.14 | 0.40 |
| **Sex, male%** | 20 (71.4%) | 42 (56.0%) | 16 (43.2%) | 0 (0.0%) | 0.15 | **0.02** | 0.20 |
| **Disease duration, months** | 50.9 (36.9) | 34.9 (24.4) | 42.2 (30.3) | 5 (%) | **0.04** | 0.33 | 0.28 |
| **Clinical phenotypes** | n=28 | n=75 | n=37 | 0 (0.0%) | 1.00 | 0.45 | 0.38 |
| ALS | 24 (85.7%) | 64 (85.3%) | 34 (91.9%) |  |  |  |  |
| ALS-FTD | 4 (14.3%) | 11 (14.7%) | 3 (8.1%) |  |  |  |  |
| **Symptom onset site** | n=28 | n=72 | n=34 | 6 (%) | 0.77 | 0.18 | 0.20 |
| Bulbar | 11 (39.3%) | 26 (36.1%) | 8 (23.5%) |  |  |  |  |
| Non-bulbar | 17 (60.7%) | 46 (63.9%) | 26 (76.5%) |  |  |  |  |
| **C9orf72** | n=26 | n=54 | n=32 | 28 (%) | 0.10 | 0.12 | 0.97 |
| Positive | 3 (11.5%) | 15 (27.8%) | 9 (28.1%) |  |  |  |  |
| Negative | 23 (88.5%) | 39 (72.2%) | 23 (71.9%) |  |  |  |  |
| **SuStaln stage** | 0 (0.0) | 5.5 (4.8) | 7.4 (5.0) | 0 (0.0%) | - | - | **0.03** |

**Note.** Continuous variables are presented as mean (standard deviation), and compared with two-sample t-test or Mann-Whitney test. Categorical variables are presented as number (percentage), and compared with Chi-square test or Fisher's exact test. Missing data indicates the percentage of individuals with missing data. P-values < 0.05 are bolded.

### Transcriptomic dynamics of subtypes

The transcriptomic dynamic trajectories of the two subtypes were constructed, each showing a distinct sequence of module involvement throughout transcriptomic progression. Each individual was assigned a SuStaIn stage within their subtype. The SuStaIn stage showed a significant correlation with z-scored MEs of each module (Supplementary Fig. 5), indicating their relevance in the transcriptomic progression. SuStaIn stage effectively reflected the severity of gene expression dysregulation. We investigated the relationship between SuStaIn stages and clinical profiles. Only in the Synapse/RNA-Metabolism subtype, SuStaIn stage was negatively correlated with age at death and symptom onset (Fig. 4a). We also compared the magnitude of SuStaIn stages across various subgroups (Fig. 4b). Females exhibited higher SuStaIn stages compared to males. Individuals carrying pathogenic variants in *C9orf72* had significantly higher SuStaIn stages than non-carriers. Moreover, Synapse/RNA-Metabolism subtype showed higher SuStaIn stages than Immune/Apoptosis/Proteostasis subtype. Fig. 4c illustrates the distribution of individuals assigned to each SuStaIn stage and their corresponding cell-type proportions, indicating that as SuStaIn stages advance, neuronal proportions decrease while microglial proportions increase. Further correlation analysis between SuStaIn stage and estimated cellular proportions revealed that within each subtype, stage was significantly negatively correlated with neuron proportion and positively correlated with microglia proportion (Fig. 4d).

**Fig. 4.**
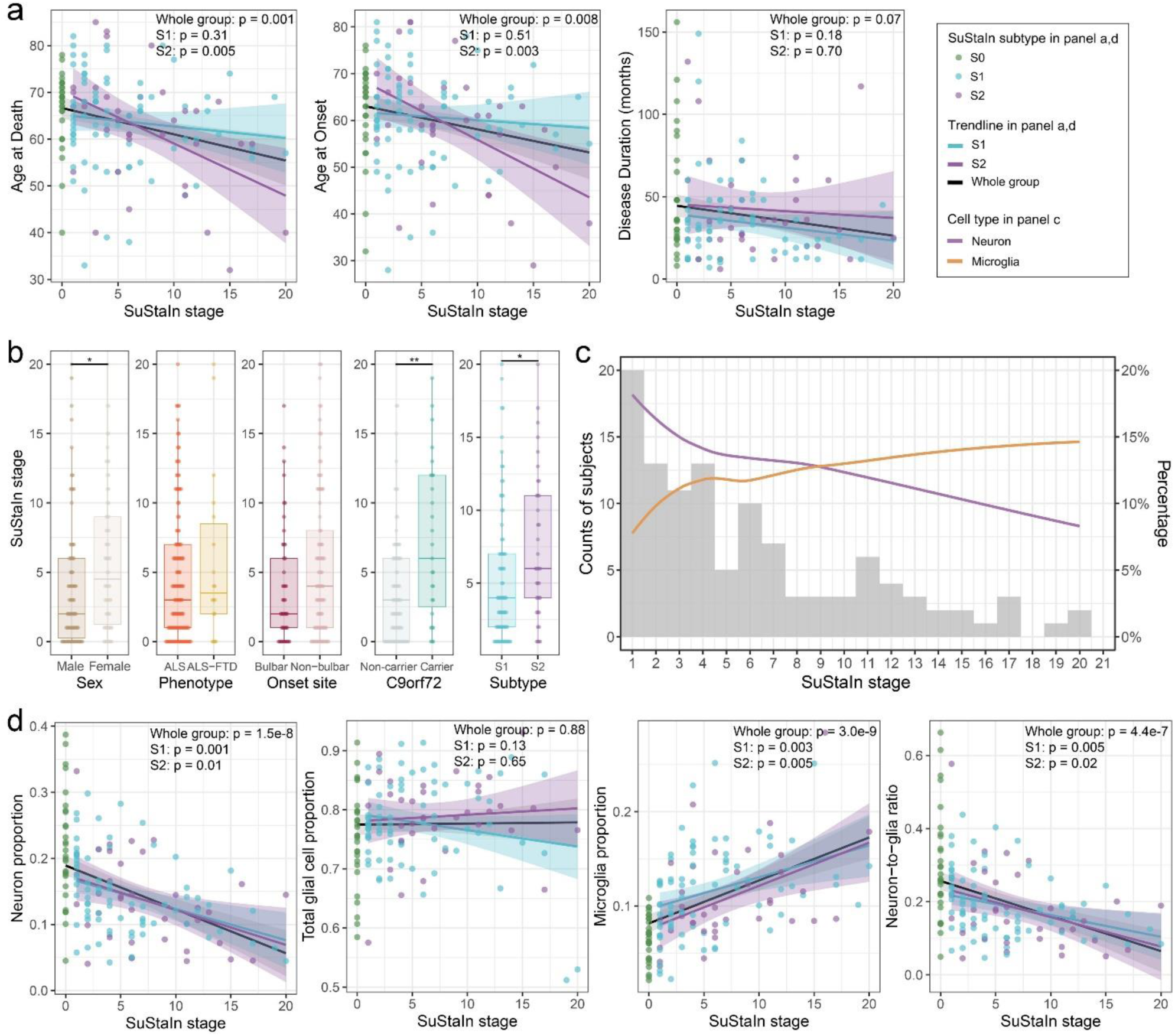
Description of SuStaIn progression trajectory. **a** SuStaIn stage correlated with age at death and symptom onset. **b** Boxplots comparing SuStaIn stage across various demographic and clinical subgroups: sex, phenotype, onset site, *C9orf72* genotype, and SuStaIn subtype. \**P* < 0.05, \*\**P* < 0.01. **c** Distribution of subjects across SuStaIn stages and cellular composition at different stages. The grey bars represent the count of subjects at each stage. The line graphs illustrate the trend lines for change of the cell-type percentages across the stages. **d** SuStaIn stage correlated with different cell-type proportions. SuStaIn = Subtype and Stage Inference; S0 = Normal-appearing group; S1 = Immune/Apoptosis/Proteostasis subtype; S2 = Synapse/RNA-Metabolism subtype.

### Validation of SuStaIn model in cervical spinal cord

The 11 Lumbar-constructed modules used for SuStaIn training, were highly preserved in cervical spinal cord samples (Fig. 5a). We projected these modules to the cervical dataset, and generated new MEs, with similar expression patterns between the two regions (Fig. 5b). The black, cyan, magenta, midnightblue, salmon, and tan modules were significantly up-regulated in ALS/ALS-FTD, while the grey60, and royalblue modules were significantly down-regulated compared to controls (Fig. 5c).

**Fig. 5.**
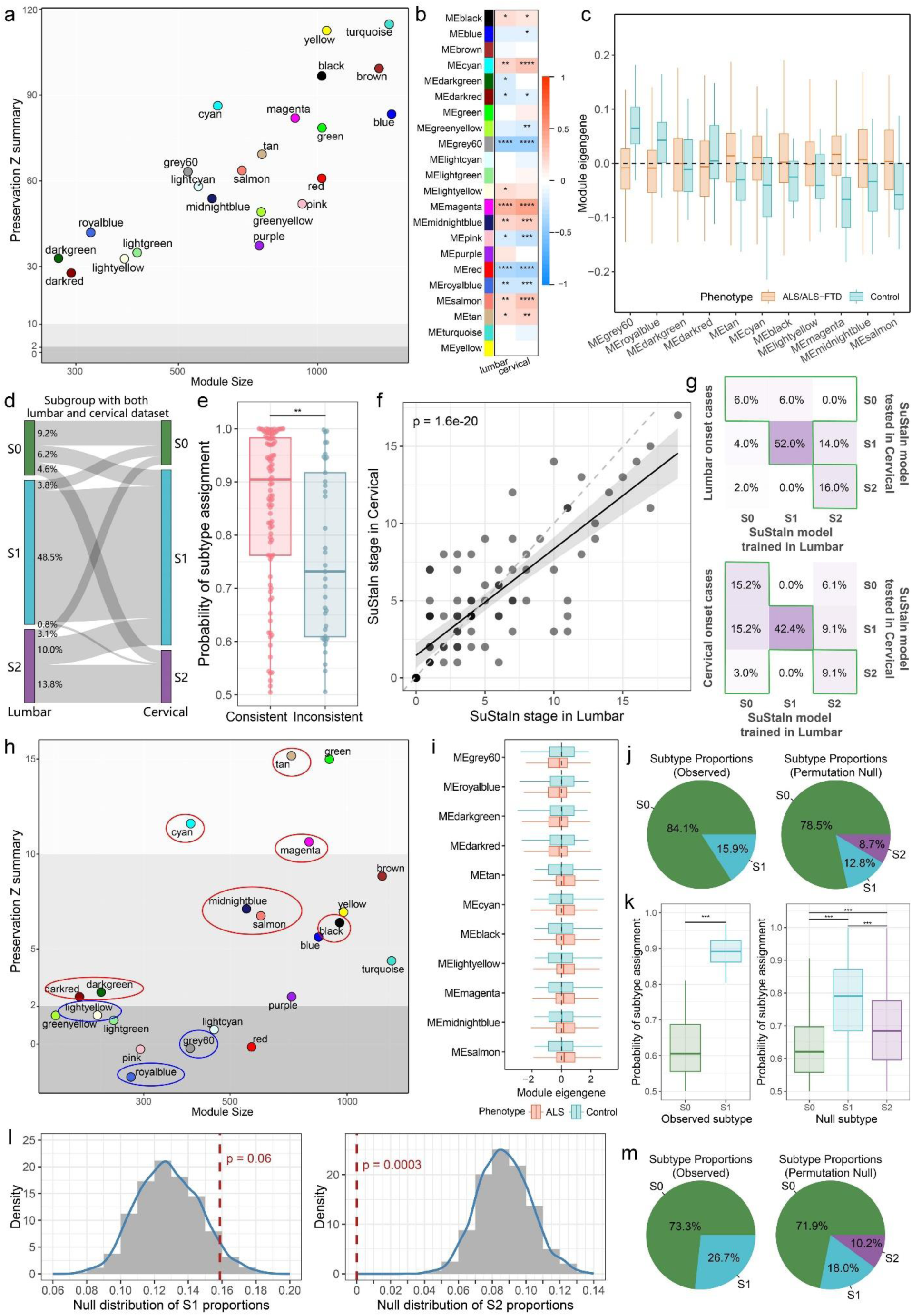
Cross-tissue Evaluation of Lumbar-trained SuStaIn model. **a** Preservation of the Lumbar-constructed modules in the cervical spinal cord. The Zsummary statistics indicate the degree of module preservation, with modules having Zsummary > 10 considered highly preserved. **b** Expression levels of the 22 modules correlated with ALS/ALS-FTD in lumbar and cervical spinal cord samples, Blue represents down-regulated and red indicates up-regulated modules in ALS/ALS-FTD relative to controls. **c** Comparisons of module eigengenes derived from cervical dataset across phenotypes. **d** Subtype assignment between lumbar and cervical datasets. **e** The subtype probability is higher in individuals with consistent subtype assignments across lumbar and cervical datasets. **f** SuStaIn stages in the lumbar are highly correlated with stages in cervical samples for individuals with consistent subtype assignments. **g** Subtype assignment consistency between lumbar and cervical datasets across different onset site groups. Green boxes indicate cases with consistent assignment when analyzing the primary onset site. **h** Preservation of the Lumbar-constructed modules in blood sample. **i** Comparisons of module eigengenes derived from blood samples across phenotypes. All 11 modules were significantly differed in ALS compared to controls. **j** Subtype proportions in observed true values and null distributions. **k** The probability of subtype assignment across SuStaIn subtypes. \*\*\**P* < 0.001. **l** Histograms display the null distribution of subtype proportions derived from permutations. The red line represents the observed subtype proportions. **m** Subtype proportions in observed and null distributions after filtering for assignment probabilities > 0.6. \**P* < 0.05, \*\**P* < 0.01, \*\*\**P* < 0.001, \*\*\*\**P* < 0.0001. SuStaIn = Subtype and Stage Inference; S0 = Normal-appearing group; S1 = Immune/Apoptosis/Proteostasis subtype; S2 = Synapse/RNA-Metabolism subtype.

We assessed subtype consistency by applying the Lumbar-trained SuStaIn model to cervical data from 130 ALS/ALS-FTD patients. There were 71.5% (93/130) remained consistent with their lumbar subtype assignments, were regarded as “subtype consistent”, while 28.5% (37/130) were considered as “subtype inconsistent” due to differing subtype assignments (Fig. 5d). The probability of subtype assignment in lumbar dataset was significantly lower in the “subtype inconsistent” group relative to the “subtype consistent” (Fig. 5e), indicating that individuals with higher assignment probability were more likely to show stable subtype assignment across datasets. In the “subtype consistent” group, the lumbar and cervical SuStaIn stages were strongly correlated (Spearman’s r=0.76, *P*<0.0001; Fig. 5f). Beyond technical limitations, we observed that subtype inconsistencies may partially be explained by their initially affected regions (Fig. 5g). Three lumbar-onset cases, which had a normal-appearing subtype in cervical samples but any subtype in lumbar samples, remained subtype-consistent; similarly, 6 cervical-onset cases with a normal-appearing subtype in lumbar samples and any subtype in cervical samples also maintained subtype consistency.

### Cross-tissue validation in peripheral blood

To evaluate the cross-tissue applicability of our model, we performed validation in peripheral blood samples from an independent ALS cohort. Of 11 Lumbar-constructed modules used for SuStaIn training, 8 were preserved in blood (Fig. 5h). Notably, core modules of Immune/Apoptosis/Proteostasis subtype including magenta, midnightblue, salmon, and cyan modules were preserved, while core modules of Synapse/RNA-Metabolism subtype, such as royalblue and lightyellow modules, along with the neuro-associated grey60 module were not, consistent with neural signature tissue specificity. Projecting the Lumbar-constructed modules to blood sample and extracting MEs showed expression change directions mirrored spinal cord, with significant ALS-control differences (Fig. 5i).

Applying the Lumbar-trained SuStaIn model to blood transcriptomic data, 63 of 397 (15.9%) ALS samples were assigned to Immune/Apoptosis/Proteostasis subtype (Fig. 5j), others to normal-appearing group. Synapse/RNA-metabolism subtype was not detected, and might be assigned to normal-appearing group, likely because its core modules were not preserved in blood. Moreover, the higher proportion of normal-appearing group may be due to early-stage blood sampling at the first clinical visit, when subtype-specific transcriptional signatures may not yet be fully established. The S1 assignment probability was substantially higher than S0, while all three subtypes showed uniformly lower probabilities in the null distribution (Fig. 5k), suggesting considerably weaker subtype stability under permuted conditions. Permutation testing yielded *P*=0.06, indicating only a marginal excess over the null distribution (Fig. 5l). However, after filtering samples with assignment probabilities < 0.6, the remaining samples were 59.4% in true values and 63.2% in null distributions, repeating the comparison yielded improved significance for S1 (*P*=0.0007, Fig. 5m).

### Comparison with a subtype-only model

To evaluate SuStaIn’s performance in capturing ALS transcriptomic subtypes, we compared it with a subtype-only model, that does not account for temporal dynamics. We employed a nsNMF clustering model on the same lumbar bulk RNA-seq dataset (19,614 genes, 171 samples). Since SuStaIn identified two transcriptomic subtypes, a rank of 2 was chosen for the nsNMF model.

Principal component analysis (PCA) revealed distinct clustering patterns between two NMF-derived subtypes (Fig. 6a). To select representative genes for each subtype, genes were ranked by their contributions to the identified subtypes. We selected the top 2,000 genes for each subtype, and found 947 common and 1,053 unique genes (Fig. 6b). We conducted functional enrichment analysis on the unique and common genes for biological annotations of the two NMF-derived subtypes (Fig. 6c). Both subtypes involved a majority of biodomains, but with distinct patterns. One subtype demonstrated unique involvement in “Immune System” and “Oxidative Stress”, along with relatively high contributions from “Apoptosis”, “RNA Metabolism”, “Proteostasis”, and “Circulatory System” biodomains, which termed as “Immune and RNA/Protein metabolism”. The other subtype was predominantly associated with “Synapse”, “Cognition and Behavior”, and “Ion Transport and Homeostasis” biodomains, named the “Synapse subtype”. Both NMF-derived subtypes showed involvement in “Neuron Generation”, “Myelination”, “Gliogenesis” and “Cytoskeleton” biodomains. Thus, the Immune and RNA/Protein Metabolism subtype demonstrated substantially broader pathological engagement, while the Synapse subtype remained more focused on synaptic functions.

**Fig. 6.**
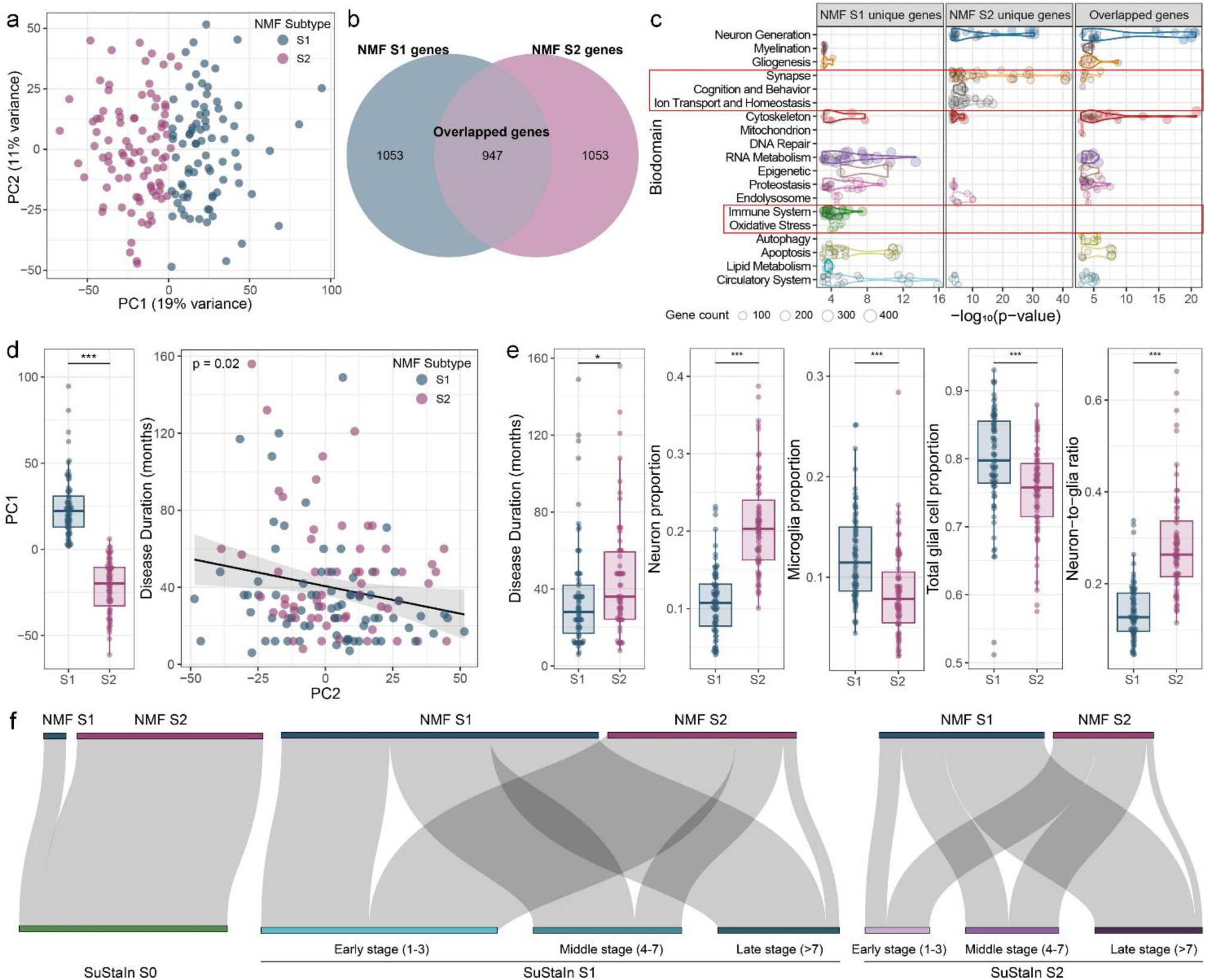
Transcriptomic subtypes identified by subtype-only model. **a** PCA shows the clustering of samples based on NMF subtypes. **b** Venn diagram illustrates the unique and shared top contributing genes of two NMF subtypes. **c** Biological annotation of subtype specific and shared genes. Each dot represents a GO term within a given biodomain. Color of the dots represent different biodomains. The x-axis indicates the significance of term enrichment, and the dot size indicates the number of genes in that GO term and in the top contributing genes of two subtypes. **d** Relationships of principal components with NMF subtyping and disease duration. **e** Distinct clinical features and cell-type proportions between NMF subtypes. **f** SuStaIn subtypes at different stages mapping to NMF subtypes. \**P* < 0.05, \*\*\**P* < 0.001. PCA = principal component analysis; NMF = non-negative matrix factorization; SuStaIn = Subtype and Stage Inference; SuStaIn S0 = Normal-appearing group; SuStaIn S1 = Immune/Apoptosis/Proteostasis subtype; SuStaIn S2 = Synapse/RNA-Metabolism subtype; NMF S1 = Immune and RNA/Protein metabolism subtype; NMF S2 = Synapse subtype.

The two NMF subtypes had an approximate ratio of 1.1:1. The first principle component (PC1) from PCA differed significantly between subtypes, and the second principle component (PC2) correlated with disease duration (Fig. 6d). The Immune and RNA/Protein metabolism subtype exhibited shorter disease duration compared to the Synapse subtype, along with reduced neuron and elevated microglia proportions (Fig. 6e, and Supplementary Table 2). NMF-derived subtypes share partial similarities with SuStaIn’s classification, but critical distinctions exist. The NMF-derived Synapse subtype was more specific, while the Immune and RNA/Protein metabolism consolidated most high-load biodomains. The SuStaIn subtypes reflected dynamic progression, with biodomain involvement expanding as the SuStaIn stages advanced. As the disease progressed to middle and late stages, the number of biodomains involved increased. These individuals, with multi-biodomain involvement, were more likely to be classified into the NMF’s Immune and RNA/Protein Metabolism subtype (Fig. 6f, and Supplementary Fig. 6).

### Potential driver genes of subtyping and staging

We identified core modules and their hub genes strongly correlated with both MEs and SuStaIn subtypes, suggesting their potential role in subtyping. Core modules of the Immune/Apoptosis/Proteostasis subtype include magenta, midnightblue, salmon, and cyan, while those of the Synapse/RNA-Metabolism subtype are royalblue, black, and lightyellow. Each module yielded a set of hub genes, which are likely drivers of subtyping (Supplementary Fig. 7a, b, Data file S3). We then additionally identified hub genes correlated with SuStaIn stages (Supplementary Fig. 7c, and Data file S3), suggesting their potential as critical drivers of transcriptomic staging. Hub genes for subtyping and staging largely overlapped (Fig. 7a).

**Fig. 7.**
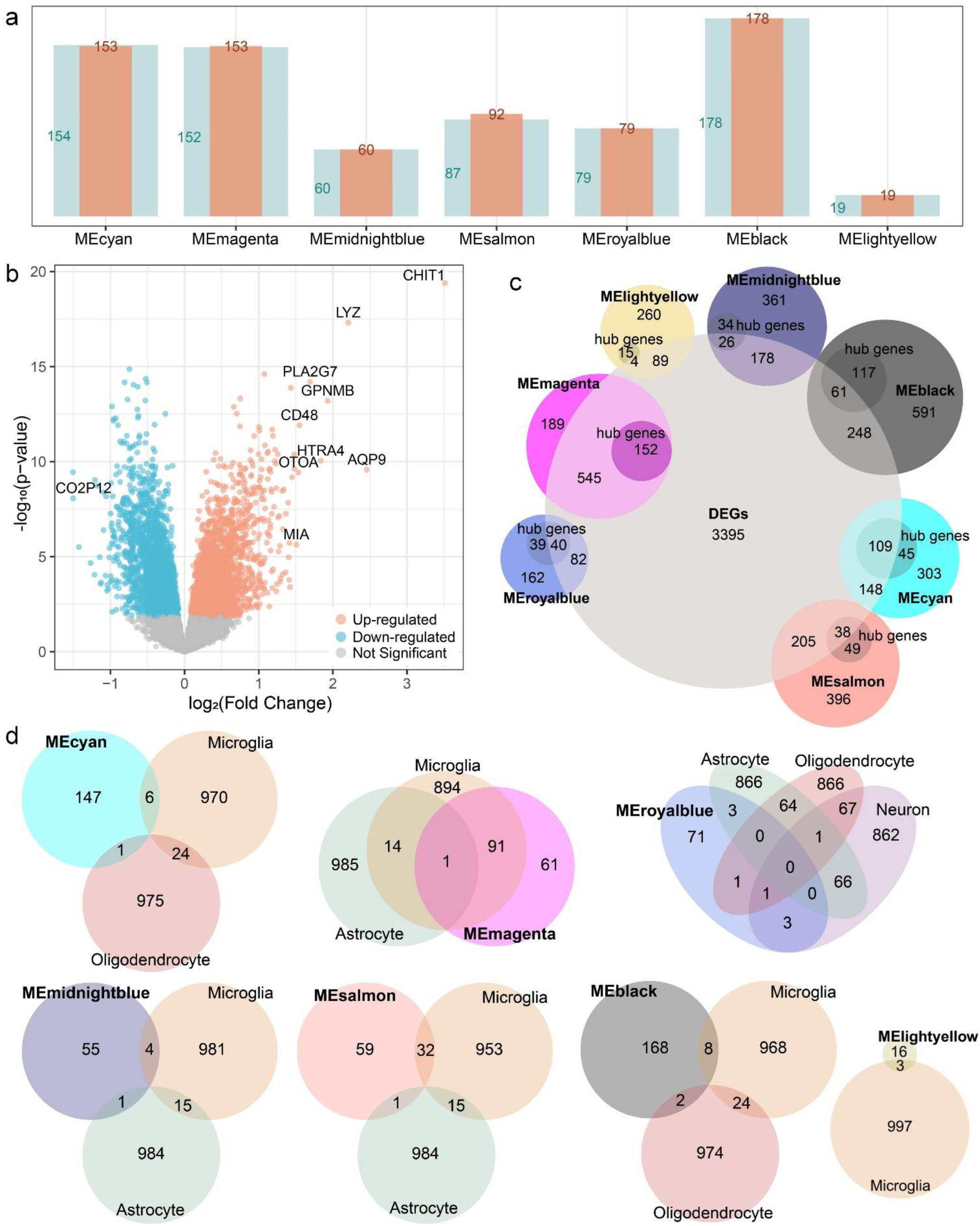
Hub genes within core modules associated with subtyping and staging. **a** Overlap between hub genes associated with subtyping and staging in core modules. The light blue bars represent hub genes associated with subtyping, and the orange bars represent hub genes associated with staging. The overlap indicates a significant intersection between the two sets of genes in each module, with the numbers adjacent to each bar corresponding to the count of hub genes within that module. **b** Volcano plot showings DEGs identified in ALS/ALS-FTD relative to controls. **c** Overlap of DEGs with core modules. For each module, the outer circle represents the total number of genes in the module, while the inner circle represents the hub genes of that module. **d** Venn diagrams illustrating the overlap between hub genes and cell type-specific gene markers across the core modules. DEG = differentially expressed gene.

Differential expression analysis on the lumbar RNA-seq data, identified 5,429 differentially expressed genes (DEGs) that were significantly up- or down-regulated in ALS/ALS-FTD relative to controls (Fig. 7b, and Data file S4). Notably, all hub genes within the magenta module were DEGs, while the other core modules contained hub genes that were not detected in traditional differential expression analyses (Fig. 7c).

To explore whether hub genes exhibit cell type-specific expression, we conducted overlap analysis with cell type-specific marker genes (Fig. 7d, Supplementary Fig. 8). Hub genes within magenta and salmon modules were predominantly enriched in microglia (permutation test *P*<0.0001), supporting their role in mediating immune responses within central nervous system (CNS). In contrast, the royalblue module that associated with “Synapse” and “Cognition and Behavior”, showed a stronger association with neurons, astrocytes, and oligodendrocytes, suggesting a different functional profile. Other core modules associated with “RNA metabolism” and “Proteostasis” biodomains did not show specific enrichment in these cell types, which might suggest broader systemic involvement beyond CNS.

### Identification of potential subtype-specific therapeutic agents

Text mining performed by Coremine Medical software using the names of top hub genes as search terms in co-occurrence analysis, and identified potential drugs associated with these genes (Fig. 8a). We found that top hub genes such as “*S100A11*”, “*GPX1*”, “*TYROBP*”, and “*NAGA*” within the immune-related magenta module, were linked to anti-inflammatory agents including recombinant interleukin-6, catechin and cysteine, capable of modulating immune pathway abnormalities. Hub genes within proteostasis-related cyan module, were associated with tyrosine, cysteine and interferon-gamma, suggesting their integrated roles in regulating proteostatic balance. Given that these two core modules characterize the Immune/Apoptosis/Proteostasis subtype, their corresponding therapeutic agents represent potential candidates for early-stage treatment in this subtype. Similarly, lysine, buformin and verteporfin were retrieved as agents capable of directly or indirectly modulating RNA homeostasis. Moreover, the thiamine, kynurenic acid, catechin and tyrosine were repeatedly co-cited with hub genes such as “*KATNB1*”, “*TMEM63B*”, “*FRS3*” and “*MAPK8IP3*” within the synaptic-related royalblue module, underscoring their candidacy as neuroprotective therapeutic targets. Thus, these therapeutic agents are potential candidates for the Synapse/RNA-Metabolism subtype. Furthermore, by screening the DrugBank database, we identified potential drugs targeting the hub genes of each subtype’s core modules. These drug candidates may help prioritize subtype-specific therapeutic strategies (Data file S5).

**Fig. 8.**
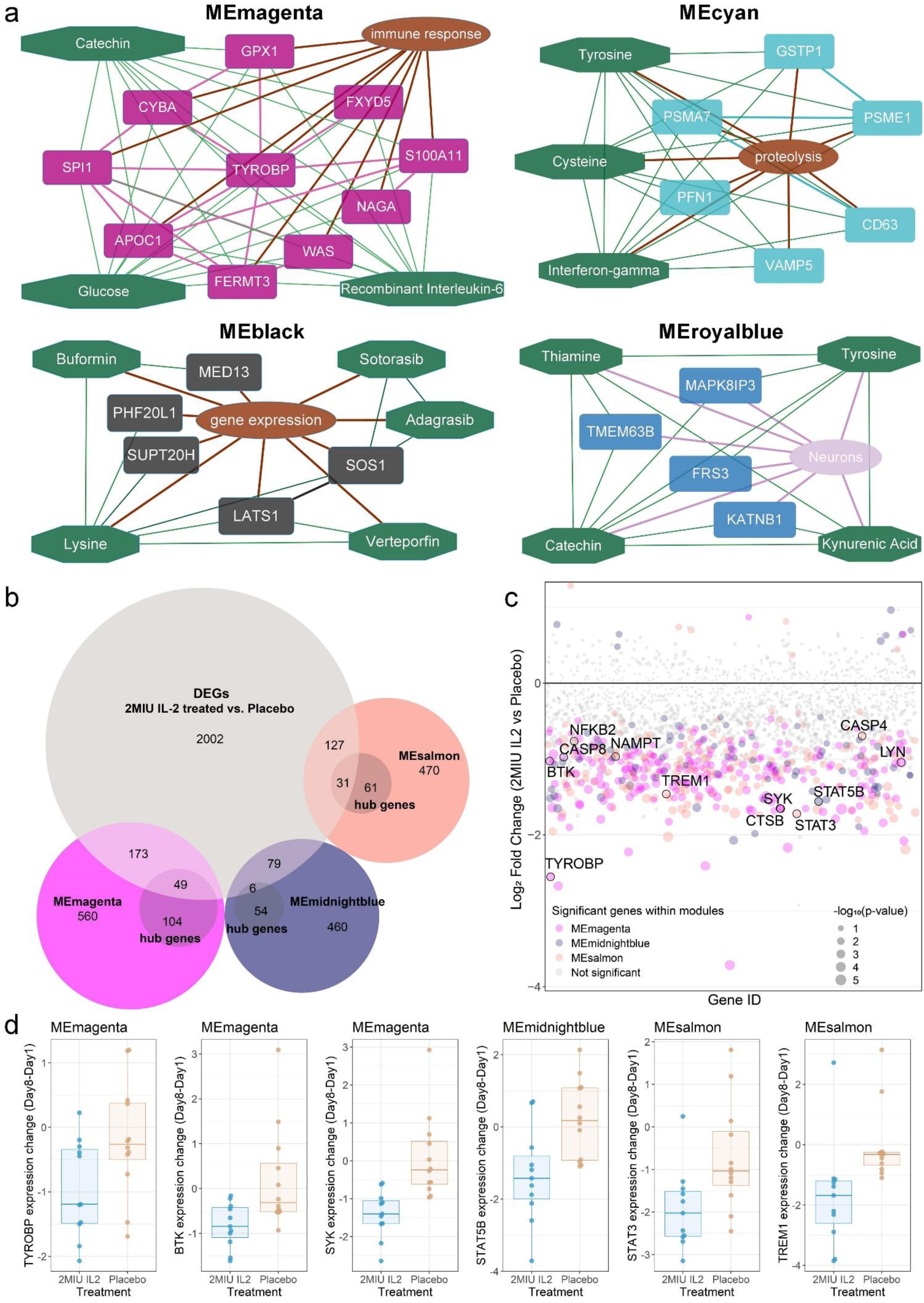
Transcriptomic response to IL-2 treatment. **a** Identification of potential drugs for top hub genes within core modules. **b** Overlap of DEGs resulting from the comparisons between treatment and placebo groups with core WGCNA modules. For each module, the outer circle represents the total number of genes in the module, while the inner circle represents the hub genes of that module. **c** Manhattan plot showing the DEGs mapping to immune-related WGCNA modules. Annotated genes indicate potential targets supported by existing literature. **d** Significant transcriptomic change from baseline (Day1) to end of first cycle of IL-2 treatment (Day8), showing several known therapeutic targets in neurodegenerative diseases. IL-2 = interleukin-2; DEG = differentially expressed gene; WGCNA = weighted gene co-expression network analysis.

### Transcriptomic responses to potential therapeutic agents

Notably, IL-2 (Aldesleukin), which targets genes (e.g., *IL2RB* and *IL2RG*) within immune-related midnightblue module, is currently under clinical investigation for ALS treatment^41^. Our reanalysis of Giovannelli et al.’s transcriptomics data revealed that a cycle of IL-2 treatment induced substantial genome-wide changes in ALS patients. Comparative analysis between the 2 million international units (MIU) IL-2 treatment group and placebo controls at day 8 identified 2,467 DEGs (uncorrected *P*<0.05). Notably, we observed significant overlap between these treatment-responsive genes and our predefined immune-related modules (Fig. 8b, c). Permutation tests (Supplementary Fig. 9) revealed that a significant fraction of each module’s genes were DEGs (*P*<0.0001): the magenta module showed significant enrichment for 222 DEGs (25.1%), including 49 hub genes representing 5.5% of module; and the salmon module contained 158 overlapping DEGs (22.9%), including 31 hub genes (4.5%). In contrast, the midnightblue module, which lacked significant enrichment for microglia markers, showed a non-significant overlap with 85 DEGs (14.2%) and 6 hub genes (1.0%) (*P*>0.05). Notably, several identified hub genes represent known therapeutic targets in neurodegenerative diseases, and their expression was significantly downregulated following IL-2 administration (Fig. 8d).

## Discussion

We employed a transcriptomic SuStaIn model to unravel temporal dynamics of distinct transcriptomic subtypes in ALS. Using post-mortem bulk RNA-seq data from spinal cord, we identified two distinct transcriptomic patterns: an Immune/Apoptosis/Proteostasis subtype, and a Synapse/RNA-Metabolism subtype. These subtypes differed in clinical characteristics, as well as neuron and microglia proportions. Moreover, the SuStaIn stages reconstructed subtype-specific temporal transcriptomic trajectories, aligning with neuronal loss and microglial activation. Our findings provided new insights into ALS temporal transcriptomic heterogeneity, and underscored potential utility for identifying therapeutic targets and facilitating patient stratification in precision medicine. More broadly, this approach demonstrates the potential to infer temporal dynamics of transcriptome from cross-sectional human tissues data.

Extensive research implicated diverse molecular mechanisms in ALS heterogeneity^1,32^. We constructed co-expressed modules that captured most proposed molecular pathways, with each module representing a specific or a group of relevant molecular pathways. Our findings are consistent with previous reported up-regulation of genes linked to inflammation and immune response, apoptosis, oxidative damage, RNA Metabolism, and mitochondrial function, alongside down-regulation of genes involved in synaptic transmission, myelination, and cytoskeletal organization^5,34,42,43^. The involvement of different molecular pathways is highly interconnected and create compounding effects that ultimately drive neurodegeneration. The activation sequence of these pathways can vary, with each potentially acting as trigger, modulator, or consequence of disease. For example, inflammation and immune activation may be secondary to protein aggregation released by damaged neurons, mitochondrial dysfunction or other stresses^44^, while it can also activated early, triggering a cascade of processes including oxidative stress, mitochondrial dysfunction, and ultimately neuronal apoptosis^45^. Such variability in predominant pathways affected among patients, necessitating precise patient stratification for delivery of tailored therapies.

Supporting evidence demonstrates ALS exhibits clinical, genetic and pathological heterogeneities, with distinct predominant molecular mechanisms likely underlying these variations. Recent research has emphasized defining subtypes to capture potential driving mechanisms of neurodegeneration^14–16,36,46^. Molecular subtypes have been identified in ALS, each was characterized by predominant involvement of distinct molecular pathways^36,46^. We identified two transcriptomic subtypes characterized by distinct sequential involvement of pathway dysregulation over molecular progression. The Immune/Apoptosis/Proteostasis subtype exhibiting early involvement of the “Immune system”, “Apoptosis”, and “Proteostasis” related pathways, while the Synapse/RNA-Metabolism subtype exhibited predominate abnormalities in “Synapse” and “RNA Metabolism” related pathways. Both subtypes exhibited transcriptomic change in shared biodomains such as “Neuron Generation”, “Myelination”, and “Gliogenesis” in a neural system module encompassing microglial/immune terms, explaining its early involvement in both subtypes. Subsequent transcriptomic changes involved mitochondrial and cytoskeletal pathways in both subtypes. Then widespread biodomain involvement emerged by middle-to-late stages. Thus, the most noticeable differences between subtypes were their initial pathway involvement during early SuStaIn stages, and the sequences in which different pathways were affected throughout the progression trajectories. These findings suggested subtype-specific mechanisms driving disease occurrence and progression, likely through distinct initially affected molecular pathways that act as triggers^44,45^. Our SuStaIn findings partly aligned with prior reports of an ALS-Glia subtype characterized by enrichment for immunological activities, microglial activation, neural cell death, and worse prognosis^36^. Similarly, our Immune/Apoptosis/Proteostasis subtype exhibited shorter survival, with its predominant immune-related biodomain correlating with survival time, suggesting early immune dysregulation may drive this subtype’s poor prognosis. This reinforces neuroimmune dysregulation as a key driver of ALS severity^47^. Eshima et al. also identified ALS-Ox subtype characterized by oxidative stress and altered synaptic signaling, and ALS-TD subtype defined by transcriptional dysregulation^36^, with overlapping molecular mechanisms between these two. Our Synapse/RNA-Metabolism subtype might provide a more comprehensive summary of them. Both two SuStaIn subtypes exhibited increased neuronal loss and microglial activation. Unlike subtype-only models captured only cross-sectional characteristics, SuStaIn provided a dynamic perspective on the distinct sequential pathways involvement across subtypes, with pathophysiological severity potentially worsening with more pathways involved. Neuronal loss and microglial activation may intensify with the progression through successive SuStaIn stages. The Immune/Apoptosis/Proteostasis subtype showed a trend towards decreased neuron proportion, while Synapse/RNA-Metabolism subtype exhibited slightly greater neuronal loss. Although this trend was not statistically significant, it may be attributed to some patients having progressed to middle or later stages with multiple molecular pathways affected. Overall, this observation highlights the potential for transcriptomic subtypes to reflect different stages of disease progression and the importance of considering the temporal dimension in ALS research.

To validate the SuStaIn model, we conducted a parallel subtype-only analysis on the same RNA-seq dataset, identifying an NMF-derived Immune and RNA/Protein metabolism subtype demonstrating widespread pathway dysregulation, and a Synapse subtype showing more selective neuronal pathway involvement. Consistent with its broader molecular pathway dysregulation, particularly in immune-related pathways, the NMF-derived Immune and RNA/Protein Metabolism subtype was associated with poorer clinical outcomes, including shorter survival, accelerated neuronal loss, and heightened microglial activation. This observation aligns with our finding that later SuStaIn stage cases were more likely to map to this NMF subtype. While these later-stage cases already affected multiple pathways, the subtype-only model cannot delineate the temporal sequence of pathway involvement. Conversely, earlier SuStaIn stages, marked by fewer affected pathways, primarily mapped to the NMF Synapse subtype, this subtype may represent a biologically earlier disease state in part of ALS patients. These findings supported the SuStaIn’s value in reconstructing disease trajectories beyond static subtype classification.

SuStaIn algorithm enables subtyping and staging^48,49^. We assessed the model robustness across spinal cord regions, using cervical samples as a test dataset and revealed a 71.5% consistency in subtype assignment between two regions. Despite model limitations, inconsistencies may partially due to onset sites among patients. Specifically, some patients exhibited transcriptomic features of one subtype in the onset region (lumbar or cervical), while the other region appeared normal and unclassifiable at the current disease stage. As disease progresses, these initially normal-appearing regions might evolve to match the affected region’s subtype. Though whether the regions would become consistent requires further investigation, cross-region consistency supports model reliability for ALS subtyping. We further evaluated model’s applicability to a clinically accessible biospecimen by applying the Lumbar-trained SuStaIn model to peripheral blood transcriptomic data. SuStaIn identified the Immune/Apoptosis/Proteostasis subtype in blood samples, consistent with previous studies that also clearly distinguished an immune subtype^42^. This suggests that ALS may reflect systemic immune dysregulation, and blood-based profiling could enable early detection of the immune subtype, potentially informing treatments such as IL-2, which is already under clinical investigation in ALS^41^. However, the Synapse/RNA-metabolism subtype was not recovered in blood, likely due to low cross-tissue conservation of modules related to nervous system, indicating that the core transcriptional signature of this subtype is largely CNS-specific and not fully captured in circulation system. These cases may have been assigned to the normal-appearing group. Therefore, validating consistency between CNS and blood-based subtyping in the same cohort, could enable subtype identification in living patients for stratified management. Future work should also incorporate longitudinal sampling to track blood transcriptional signatures evolution and whether more patients transition from normal-appearing group into defined subtype trajectories at later disease stages.

Distinct transcriptomic subtypes in ALS were characterized by specific core modules, within which we identified hub genes that may act as critical drivers of both subtyping and staging in the disease (Data file S3). For the Immune/Apoptosis/Proteostasis subtype, core modules highlighted several hub genes associated with immune responses and microglial activation (*TYROBP*, *GRN*, *TREM2*, *STAT6*, *FCGRT*, etc.), and those linked to apoptosis (*TNFRSF1B*, *CASP4*, *CDKN1A*, etc.), along with *GPX1* gene involved in responding to oxidative stress. Additionally, this subtype featured a “Proteostasis” module with hub genes involved in protein degradation (*PSME1*, *PSMA7*, etc.) and some genes encoding ribosomal proteins (*RPS2*, *RPL36*, *RPL8*, etc.) that contribute to protein synthesis. All of these genes were upregulated in individuals assigned to this subtype. For the Synapse/RNA-Metabolism subtype, we identified hub genes that associated with synaptic plasticity and signaling (*CLSTN1*, *SCAMP5*, *IQCJ-SCHIP1*, *RASGRF1*, etc.). Identifying hub genes specific to each subtype may potentially guide targeted therapeutic strategies^50^. Numerous therapeutic drugs targeting mechanisms, including oxidative stress, excitotoxicity, neuroinflammation, autophagy, energy metabolism, etc., are currently under clinical investigation^51^. To identify more targeted therapeutic strategies, we employed a text-mining approach to screen for potential drugs specifically targeting the core modules that define each molecular subtype. Given that these core modules represent the earliest dysregulated biological domains in each subtype, early intervention with module-targeting drugs holds promise for achieving precise treatment. For instance, the prominent role of immune-related pathways in poorer prognosis has motivated clinical translation. The IL-2 treatment targeting the regulatory T-cells, reduced mortality risk predominantly in patients with low CSF-pNFH levels^41^, alongside measurable suppression of inflammatory cascades in treated ALS cohorts^40^. Notably, these effects were particularly pronounced effects in the Immune/Apoptosis/Proteostasis subtype, suggesting that early immunomodulatory interventions (e.g., anti-neuroinflammatory agents) may be especially beneficial for this subset of patients. These findings established molecular subtyping as a critical framework for advancing precision medicine, enabling subtype specific therapeutic strategies.

Several limitations should be considered in future work. Firstly, the lack of clinical profiles including motor and neuropsychological assessments limits the ability to fully characterize clinical features of distinct subtypes, highlighting requirements of larger cohorts with comprehensive clinical assessments for further validation. Secondly, SuStaIn model’s assumption of a monotonic progression trajectory may not fully capture the potential fluctuations in transcriptomic dynamics that might occur due to treatment effects or other factors, suggesting a need for more flexible modeling approaches that can accommodate the possible fluctuations or reversals in biomarker changes in future work. Moreover, bulk RNA-seq data might be inherently limited by cellular heterogeneity, however, it is critical to note that understanding disease heterogeneity necessitates a substantial dataset to effectively train the data-driven SuStaIn model. This requirement is currently cost-prohibitive when using snRNA-seq. Furthermore, post-mortem human tissues reflect end-stage pathology, they underscore the need to study earlier disease phases. This necessitates the collection of longitudinal biofluid samples (e.g., CSF, blood), which would both validate our framework and capture dynamic molecular changes throughout disease progression.

## Conclusions

In conclusion, we used bulk RNA-seq data from human post-mortem tissue to unravel the temporal transcriptomic signatures within individuals with ALS, disentangling the disease heterogeneities by transcriptomic subtyping and staging. We identified one subtype characterized by predominance in immune system and apoptosis-related pathways, and another primarily associated with synaptic pathways, with subsequent involvement of other molecular pathways as the disease progresses. Identification of subtype-specific transcriptomic signatures supporting the existence of transcriptomic drivers of disease heterogeneity, and also enable the prediction of the dynamic progression across distinct subtypes. This knowledge might pave the way for precisely tailored strategies in diagnosis, management, and potential treatments.

## Data availability

All RNA-seq data from NYGC ALS Consortium can be accessed via the National Center for Biotechnology Information’s Gene Expression Omnibus (GEO) database (GSE137810, GSE124439, GSE116622 and GSE153960). Two snRNA-seq datasets used are available in the GEO database (GSE243077 and GSE205521). The brain cell-type specific gene lists can be accessed at doi: 10.1038/s41598-018-27293-5. To request access to new and ongoing data generated by the NYGC ALS Consortium and for samples provided through the Target ALS Postmortem Core, complete a genetic data request form at. The RNA-seq data from peripheral blood samples is available in the GEO database (GSE112681). The microarray gene expression profiling of the IL-2 clinical trial is available in the GEO database (GSE163560). Other datasets used and/or analyzed during the current study are available from the corresponding author on reasonable request and approval from the Penn Neurodegenerative Data Sharing Committee. Requests may be submitted using a webform request: https://www.pennbindlab.com/data-sharing.

## Code availability

All packages and software described in the Methods section were used according to standard protocols. Software packages used herein include featureCounts v2.0.2 (https://subread.sourceforge.net/), GTEx gene model pipeline (https://github.com/broadinstitute/gtex-pipeline), Limma package (https://bioconductor.org/packages/release/bioc/html/limma.html), DESeq2 (https://bioconductor.org/packages/devel/bioc/vignettes/DESeq2/inst/doc/DESeq2.html), WGCNA (https://cran.r-project.org/web/packages/WGCNA/index.html), gprofiler2 (https://cran.r-project.org/web/packages/gprofiler2/vignettes/gprofiler2.html), SuStaIn (https://github.com/ucl-pond/pySuStaIn), Scaden (https://github.com/KevinMenden/scaden), NMF package (https://cran.r-project.org/web/packages/NMF/index.html), Python v3.8.3 (https://www.python.org/), and R v4.3.1 (https://www.r-project.org/).

## Acknowledgements

This work was supported by NIH funding (P01AG066597, P30AG072979, RF1NS145263, U24AG041689, U54AG052427), Penn Institute on Aging and DeCrane Fund for Primary Progressive Aphasia. The NYGC ALS Consortium was supported by ALS Association Grant 19-SI-459 and by the Tow Foundation.

## Author contributions

T.S., and C.T.M. conceived and designed the study. H.P. contributed to data collection. B.S., P.P.K., and E.B.L. contributed to RNA-seq data analysis and cell-type deconvolution. V.V.D. contributed to genetic and clinical data analysis. T.S. and C.T.M. wrote and reviewed the paper. All authors edited the paper.

## References

1 Taylor, J. P., Brown, R. H., Jr. & Cleveland, D. W. Decoding ALS: from genes to mechanism. Nature 539, 197–206 (2016). 10.1038/nature20413

2 Wood, A., Gurfinkel, Y., Polain, N., Lamont, W. & Lyn Rea, S. Molecular Mechanisms Underlying TDP-43 Pathology in Cellular and Animal Models of ALS and FTLD. Int J Mol Sci 22 (2021). 10.3390/ijms22094705

3 Brown, A. L. et al. TDP-43 loss and ALS-risk SNPs drive mis-splicing and depletion of UNC13A. Nature 603, 131–137 (2022). 10.1038/s41586-022-04436-3

4 Liu, E. Y. et al. Loss of Nuclear TDP-43 Is Associated with Decondensation of LINE Retrotransposons. Cell Rep 27, 1409–1421 e1406 (2019). 10.1016/j.celrep.2019.04.003

5 D’Erchia, A. M. et al. Massive transcriptome sequencing of human spinal cord tissues provides new insights into motor neuron degeneration in ALS. Sci Rep 7, 10046 (2017). 10.1038/s41598-017-10488-7

6 Amin, A., Perera, N. D., Beart, P. M., Turner, B. J. & Shabanpoor, F. Amyotrophic Lateral Sclerosis and Autophagy: Dysfunction and Therapeutic Targeting. Cells 9 (2020). 10.3390/cells9112413

7 Humphrey, J. et al. Integrative transcriptomic analysis of the amyotrophic lateral sclerosis spinal cord implicates glial activation and suggests new risk genes. Nat Neurosci 26, 150–162 (2023). 10.1038/s41593-022-01205-3

8 Hasan, R. et al. Transcriptomic analysis of frontotemporal lobar degeneration with TDP-43 pathology reveals cellular alterations across multiple brain regions. Acta Neuropathol 143, 383–401 (2022). 10.1007/s00401-021-02399-9

9 Li, J. et al. Divergent single cell transcriptome and epigenome alterations in ALS and FTD patients with C9orf72 mutation. Nat Commun 14, 5714 (2023). 10.1038/s41467-023-41033-y

10 Yazar, V. et al. Impaired ATF3 signaling involves SNAP25 in SOD1 mutant ALS patients. Sci Rep 13, 12019 (2023). 10.1038/s41598-023-38684-8

11 Fonteijn, H. M. et al. An event-based model for disease progression and its application in familial Alzheimer’s disease and Huntington’s disease. Neuroimage 60, 1880–1889 (2012). 10.1016/j.neuroimage.2012.01.062

12 Eshaghi, A. et al. Progression of regional grey matter atrophy in multiple sclerosis. Brain 141, 1665–1677 (2018). 10.1093/brain/awy088

13 Oxtoby, N. P. et al. Data-driven models of dominantly-inherited Alzheimer’s disease progression. Brain 141, 1529–1544 (2018). 10.1093/brain/awy050

14 Whitwell, J. L. et al. Distinct anatomical subtypes of the behavioural variant of frontotemporal dementia: a cluster analysis study. Brain 132, 2932–2946 (2009). 10.1093/brain/awp232

15 Noh, Y. et al. Anatomical heterogeneity of Alzheimer disease: based on cortical thickness on MRIs. Neurology 83, 1936–1944 (2014). 10.1212/WNL.0000000000001003

16 Bede, P., Murad, A., Lope, J., Hardiman, O. & Chang, K. M. Clusters of anatomical disease-burden patterns in ALS: a data-driven approach confirms radiological subtypes. J Neurol 269, 4404–4413 (2022). 10.1007/s00415-022-11081-3

17 Young, A. L. et al. Uncovering the heterogeneity and temporal complexity of neurodegenerative diseases with Subtype and Stage Inference. Nat Commun 9, 4273 (2018). 10.1038/s41467-018-05892-0

18 Young, A. L. et al. Data-driven neuropathological staging and subtyping of TDP-43 proteinopathies. Brain 146, 2975–2988 (2023). 10.1093/brain/awad145

19 Shen, T. et al. Novel data-driven subtypes and stages of brain atrophy in the ALS-FTD spectrum. Transl Neurodegener 12, 57 (2023). 10.1186/s40035-023-00389-3

20 Tam, O. H. et al. Postmortem Cortex Samples Identify Distinct Molecular Subtypes of ALS: Retrotransposon Activation, Oxidative Stress, and Activated Glia. Cell Rep 29, 1164–1177 e1165 (2019). 10.1016/j.celrep.2019.09.066

21 Liao, Y., Smyth, G. K. & Shi, W. featureCounts: an efficient general purpose program for assigning sequence reads to genomic features. Bioinformatics 30, 923–930 (2014). 10.1093/bioinformatics/btt656

22 Love, M. I., Huber, W. & Anders, S. Moderated estimation of fold change and dispersion for RNA-seq data with DESeq2. Genome Biol 15, 550 (2014). 10.1186/s13059-014-0550-8

23 Menden, K. et al. Deep learning-based cell composition analysis from tissue expression profiles. Sci Adv 6, eaba2619 (2020). 10.1126/sciadv.aba2619

24 Zhang, D. et al. Spatial transcriptomics and single-nucleus RNA sequencing reveal a transcriptomic atlas of adult human spinal cord. Elife 12 (2024). 10.7554/eLife.92046

25 Langfelder, P. & Horvath, S. WGCNA: an R package for weighted correlation network analysis. BMC Bioinformatics 9, 559 (2008). 10.1186/1471-2105-9-559

26 McKenzie, A. T. et al. Brain Cell Type Specific Gene Expression and Co-expression Network Architectures. Sci Rep 8, 8868 (2018). 10.1038/s41598-018-27293-5

27 Shen, T. et al. Disparate and shared transcriptomic signatures associated with cortical atrophy in genetic behavioral variant frontotemporal degeneration. Mol Neurodegener 20, 17 (2025). 10.1186/s13024-025-00806-3

28 Langfelder, P., Luo, R., Oldham, M. C. & Horvath, S. Is my network module preserved and reproducible? PLoS Comput Biol 7, e1001057 (2011). 10.1371/journal.pcbi.1001057

29 Kolberg, L. et al. g:Profiler-interoperable web service for functional enrichment analysis and gene identifier mapping (2023 update). Nucleic Acids Res 51, W207–W212 (2023). 10.1093/nar/gkad347

30 Reimand, J., Kull, M., Peterson, H., Hansen, J. & Vilo, J. g:Profiler--a web-based toolset for functional profiling of gene lists from large-scale experiments. Nucleic Acids Res 35, W193–200 (2007). 10.1093/nar/gkm226

31 Mol, M. O., Miedema, S. S. M., van Swieten, J. C., van Rooij, J. G. J. & Dopper, E. G. P. Molecular Pathways Involved in Frontotemporal Lobar Degeneration with TDP-43 Proteinopathy: What Can We Learn from Proteomics? Int J Mol Sci 22 (2021). 10.3390/ijms221910298

32 Le Gall, L. et al. Molecular and Cellular Mechanisms Affected in ALS. J Pers Med 10 (2020). 10.3390/jpm10030101

33 Cary, G. A. et al. Genetic and multi-omic risk assessment of Alzheimer’s disease implicates core associated biological domains. Alzheimers Dement (N Y) 10, e12461 (2024). 10.1002/trc2.12461

34 van Rheenen, W. et al. Whole blood transcriptome analysis in amyotrophic lateral sclerosis: A biomarker study. PLoS One 13, e0198874 (2018). 10.1371/journal.pone.0198874

35 Brunet, J. P., Tamayo, P., Golub, T. R. & Mesirov, J. P. Metagenes and molecular pattern discovery using matrix factorization. Proc Natl Acad Sci U S A 101, 4164–4169 (2004). 10.1073/pnas.0308531101

36 Eshima, J. et al. Molecular subtypes of ALS are associated with differences in patient prognosis. Nat Commun 14, 95 (2023). 10.1038/s41467-022-35494-w

37 Gaujoux, R. & Seoighe, C. A flexible R package for nonnegative matrix factorization. BMC Bioinformatics 11, 367 (2010). 10.1186/1471-2105-11-367

38 Jenssen, T. K., Laegreid, A., Komorowski, J. & Hovig, E. A literature network of human genes for high-throughput analysis of gene expression. Nat Genet 28, 21–28 (2001). 10.1038/ng0501-21

39 Knox, C. et al. DrugBank 6.0: the DrugBank Knowledgebase for 2024. Nucleic Acids Res 52, D1265–D1275 (2024). 10.1093/nar/gkad976

40 Giovannelli, I. et al. Amyotrophic lateral sclerosis transcriptomics reveals immunological effects of low-dose interleukin-2. Brain Commun 3, fcab141 (2021). 10.1093/braincomms/fcab141

41 Bensimon, G. et al. Efficacy and safety of low-dose IL-2 as an add-on therapy to riluzole (MIROCALS): a phase 2b, double-blind, randomised, placebo-controlled trial. Lancet 405, 1837–1850 (2025). 10.1016/S0140-6736(25)00262-4

42 Grima, N. et al. RNA sequencing of peripheral blood in amyotrophic lateral sclerosis reveals distinct molecular subtypes: Considerations for biomarker discovery. Neuropathol Appl Neurobiol 49, e12943 (2023). 10.1111/nan.12943

43 Namboori, S. C. et al. Single-cell transcriptomics identifies master regulators of neurodegeneration in SOD1 ALS iPSC-derived motor neurons. Stem Cell Reports 16, 3020–3035 (2021). 10.1016/j.stemcr.2021.10.010

44 Beland, L. C. et al. Immunity in amyotrophic lateral sclerosis: blurred lines between excessive inflammation and inefficient immune responses. Brain Commun 2, fcaa124 (2020). 10.1093/braincomms/fcaa124

45 Zhang, W., Xiao, D., Mao, Q. & Xia, H. Role of neuroinflammation in neurodegeneration development. Signal Transduct Target Ther 8, 267 (2023). 10.1038/s41392-023-01486-5

46 Morello, G. et al. Integrative multi-omic analysis identifies new drivers and pathways in molecularly distinct subtypes of ALS. Sci Rep 9, 9968 (2019). 10.1038/s41598-019-46355-w

47 Liu, J. & Wang, F. Role of Neuroinflammation in Amyotrophic Lateral Sclerosis: Cellular Mechanisms and Therapeutic Implications. Front Immunol 8, 1005 (2017). 10.3389/fimmu.2017.01005

48 Young, A. L. et al. Characterizing the Clinical Features and Atrophy Patterns of MAPT-Related Frontotemporal Dementia With Disease Progression Modeling. Neurology 97, e941–e952 (2021). 10.1212/WNL.0000000000012410

49 Vogel, J. W. et al. Four distinct trajectories of tau deposition identified in Alzheimer’s disease. Nat Med 27, 871–881 (2021). 10.1038/s41591-021-01309-6

50 Faller, K. M. E., Chaytow, H. & Gillingwater, T. H. Targeting common disease pathomechanisms to treat amyotrophic lateral sclerosis. Nat Rev Neurol 21, 86–102 (2025). 10.1038/s41582-024-01049-4

51 Tzeplaeff, L., Wilfling, S., Requardt, M. V. & Herdick, M. Current State and Future Directions in the Therapy of ALS. Cells 12 (2023). 10.3390/cells12111523

