## Supplementary tables and figures for "Unraveling temporal dynamics of the post-mortem transcriptome in amyotrophic lateral sclerosis"

**Supplementary Tables 1 and 2**

**Supplementary Figures 1 to 9**

**Legends for Data file S1 to S5**

- **Data file S1:** Genes clustered into co-expressed modules using weighted gene co-expression network analysis (WGCNA).
- **Data file S2:** Functional enrichment analysis of each module identified in Data file S1.
- **Data file S3:** Lists the hub genes within core modules that show significant associations with SuStaIn subtyping and/or staging.
- **Data file S4:** Differentially expressed genes (DEGs) in ALS and their corresponding WGCNA module assignments.
- **Data file S5:** Drugs targeting hub genes in Data file S3, retrieved from the Drugbank database.

Table S1. Clinical and technical characteristics of the study cohort.

| | Control | ALS | ALS-FTD | $P_{ALSvsControl}$ | $P_{ALS\text{-}FTDvsControl}$ | $P_{ALSvsALS\text{-}FTD}$ |
|---|---|---|---|---|---|---|
| **Donors, n** | 45 | 151 | 21 | - | - | - |
| **Sex (male%)** | 22 (48.9%) | 79 (52.3%) | 13 (61.9%) | 0.69 | 0.32 | 0.41 |
| **Age at onset (years)** | - | 60.5 (10.7) | 63.1 (11.3) | - | - | 0.20 |
| **Age at death (years)** | 63.7 (16.9) | 64.2 (10.0) | 67.4 (9.6) | 0.77 | 0.44 | 0.11 |
| **Disease duration (months)** | - | 40.2(29.2) | 31.7 (29.0) | - | - | **0.04** |
| **Symptom onset site** | - | n = 145 | n = 20 | - | - | 0.08 |
| Bulbar | - | 44 (30.3%) | 10 (50.0%) | - | - | - |
| Non-bulbar | - | 101 (69.7%) | 10 (50.0%) | - | - | - |
| **Initially affected region**[a] | - | n = 135 | n = 19 | - | - | 0.31 |
| Bulbar | - | 37 (27.4%) | 9 (47.4%) | - | - | - |
| Cervical/ Cervical+Bulbar | - | 37 (27.4%) | 3 (15.8%) | | | |
| Lumbar/Lumbar+Bulbar | - | 59 (43.7%) | 7 (36.8%) | | | |
| Cervical + Lumbar mixed | - | 2 (1.5%) | 0 (0.0%) | - | - | - |
| **Cause of death** | | | | **< 0.0001** | **< 0.0001** | 0.97 |
| Refers to ALS explicitly | 0 (0.0%) | 60 (39.7%) | 6 (28.6%) | - | - | - |
| Organ failure consistent with neurodegenerative state | 0 (0.0%) | 22 (14.6%) | 5 (23.8%) | - | - | - |
| Cardio & Cerebrovascular diseases | 18 (40.0%) | 1 (0.7%) | 0 (0.0%) | - | - | - |
| Infection | 6 (13.3%) | 6 (4.0%) | 3 (14.3%) | - | - | - |
| Malignancy | 11 (24.4%) | 0 (0.0%) | 0 (0.0%) | - | - | - |
| Organ failure due to other diseases | 8 (17.8%) | 0 (0.0%) | 0 (0.0%) | - | - | - |
| Voluntary Termination | 1 (2.2%) | 19 (12.6%) | 0 (0.0%) | - | - | - |
| Not reported | 1 (2.2%) | 43 (28.5%) | 7 (33.3%) | - | - | - |
| ***C9orf72*** | n = 13 | n = 123 | n = 18 | 0.07 | **0.01** | **<0.05** |
| Positive | 0 (0.0%) | 28 (22.8%) | 8 (44.4%) | - | - | - |
| Negative | 13 (100.0%) | 95 (77.2%) | 10 (55.6%) | - | - | - |
| **Tissues** | | | | | | |
| Lumbar spinal cord, n | 31 | 121 | 18 | - | - | - |
| $RIN_{lumbar}$ | 6.0 (0.8) | 6.7 (0.9) | 6.7 (0.8) | **<0.0001** | **0.004** | 0.93 |
| Cervical spinal cord, n | 34 | 142 | 20 | - | - | - |
| $RIN_{cervical}$ | 6.5 (0.7) | 6.8 (0.9) | 6.8 (0.6) | **0.007** | 0.05 | 0.92 |

Note. Continuous variables are presented as mean (standard deviation), and compared with two-sample t-test or Mann-Whitney test. Categorical variables are presented as number (percentage), and compared with Chi-square test or Fisher's exact test. [a] the initially affected region were based on the symptom onset site. $P < 0.05$ are bolded. RIN, RNA integrity number.

Table S2. Comparison of characteristics between NMF subtypes.

| | S1: Immune and RNA/Protein metabolism subtype | S2: Synapse subtype | $P_{S1vsS2}$ |
|---|---|---|---|
| **Subject, n** | 73 | 67 | - |
| **Age at Death, years** | 64.0 (11.3) | 63.8 (9.5) | 0.96 |
| **Age at symptom onset, years** | 61.0 (11.5) | 60.0 (10.4) | 0.77 |
| **Sex, male%** | 37 (50.7%) | 41 (61.2%) | 0.21 |
| **Disease duration, months** | 35.3 (27.5) | 45.4 (30.5) | **0.01** |
| **Clinical phenotypes** | n = 73 | n = 67 | 0.19 |
| ALS | 61 (83.6%) | 61 (91.0%) | |
| ALS-FTD | 12 (16.4%) | 6 (9.0%) | |
| **Symptom onset site** | n = 71 | n = 63 | 0.08 |
| Bulbar | 19 (26.8%) | 26 (41.3%) | |
| Non-bulbar | 52 (73.2%) | 37 (58.7%) | |
| ***C9orf72*** | n = 56 | n=56 | 0.83 |
| Positive | 14 (25.0%) | 13 (23.2%) | |
| Negative | 42 (75.0%) | 43 (76.8%) | |
| **PC1** | 24.5 (17.8) | -21.6 (15.5) | **< 0.0001** |
| **PC2** | 0.9 (21.6) | 3.3 (19.5) | 0.51 |

Note. Continuous variables are presented as mean (standard deviation), and compared with two-sample t-test or Mann-Whitney test. Categorical variables are presented as number (percentage), and compared with Chi-square test or Fisher's exact test. Missing data indicates the percentage of individuals with missing data. $P < 0.05$ are bolded. S1, Immune and RNA/Protein metabolism subtype; S2, Synapse subtype; NMF, non-negative matrix factorization; PC, principal component.

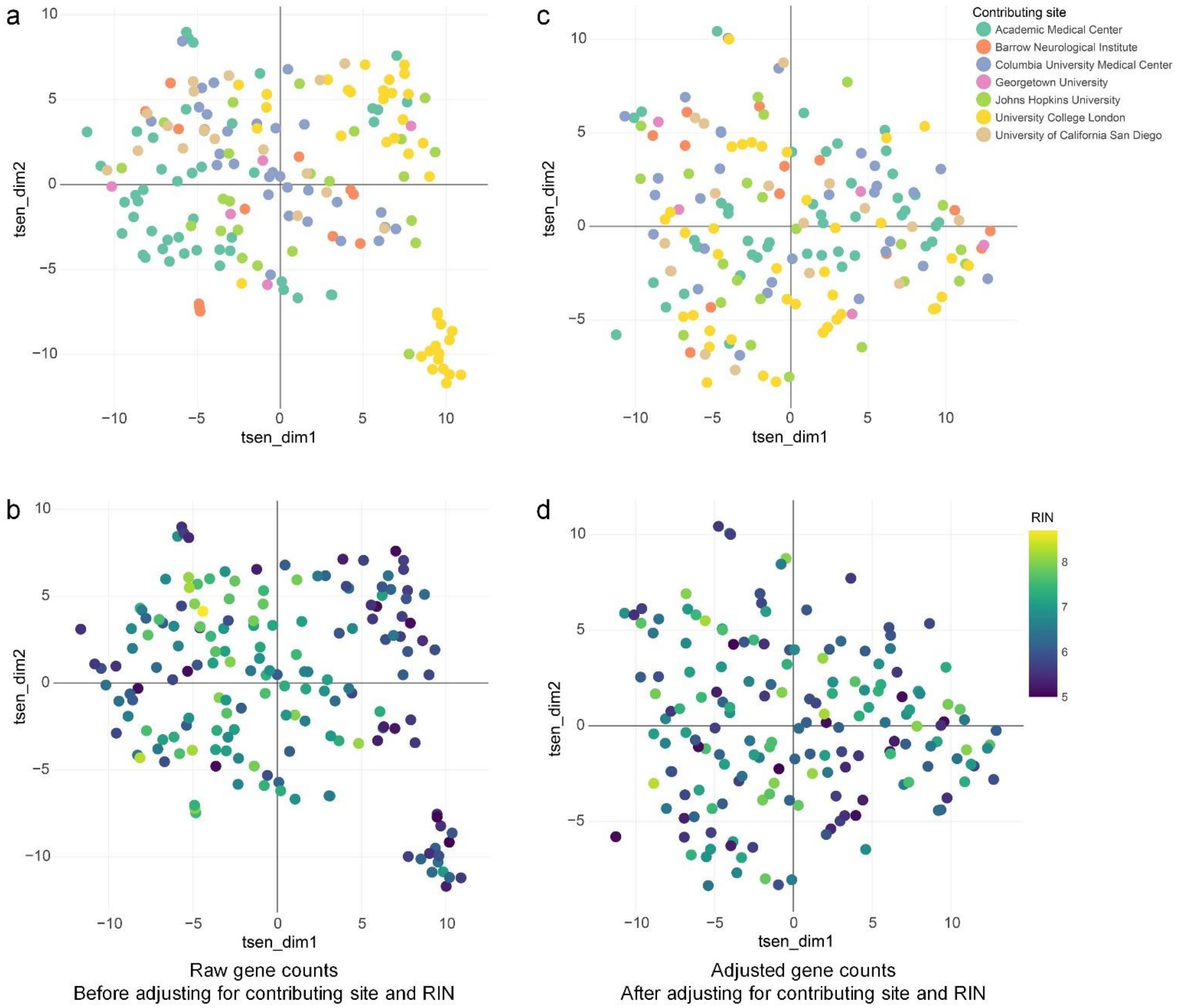


**Supplementary Fig. 1 T-distributed stochastic neighbor embedding visualization of the lumbar RNA-seq dataset.** Visualization of the raw RNA-seq data, colored by different contributing sites (**a**), or by RIN (**b**). Visualization of the RNA-seq data after adjusted for contributing site and RIN, colored by different contributing sites (**c**), or by RIN (**d**). RIN = RNA integrity number.

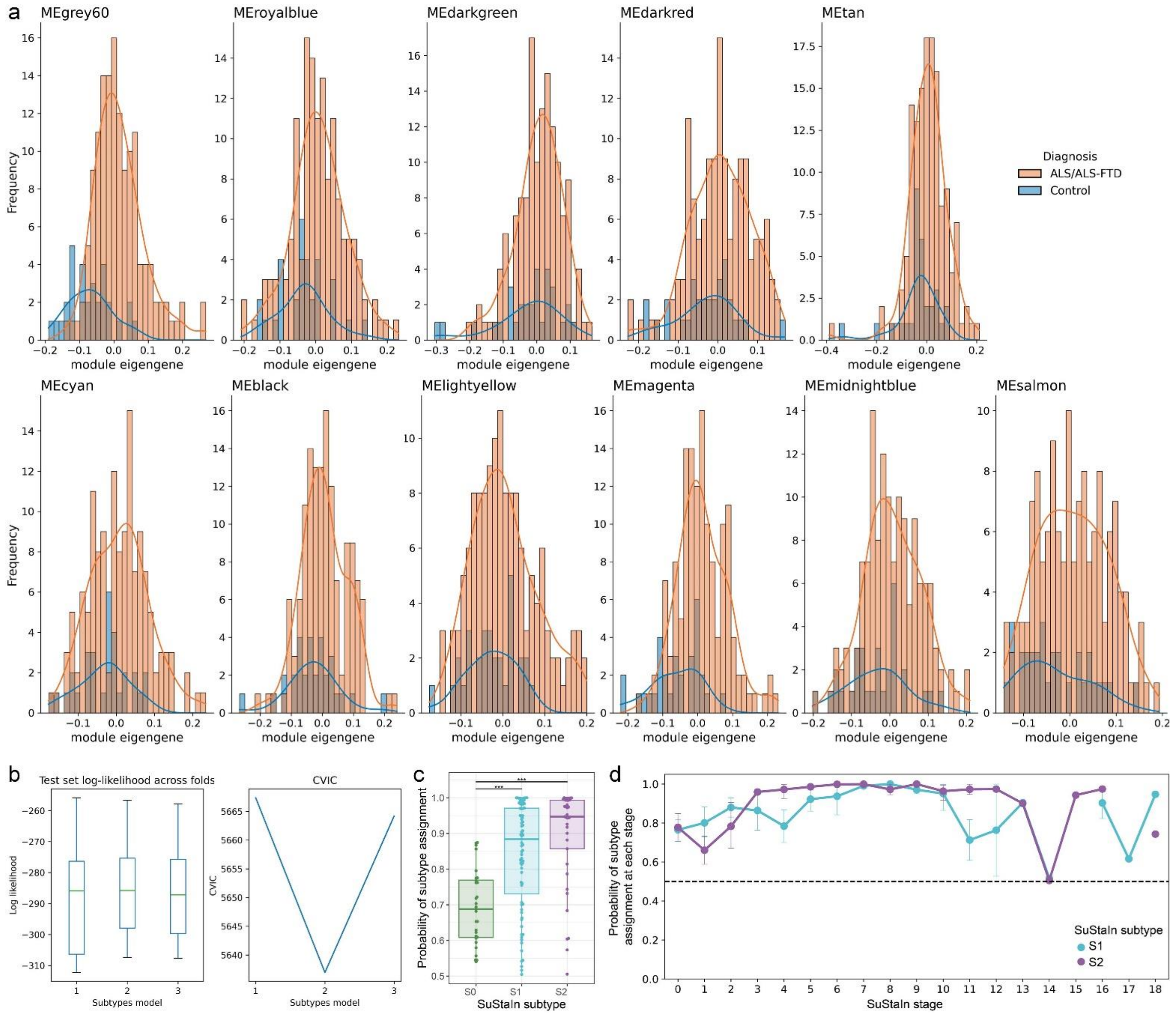


**Supplementary Fig. 2 Building and evaluating the SuStaIn model. a** Module eigengenes across clinical phenotypes for each module. **b** Cross-validation was employed by calculating the out-of-sample log-likelihood and CVIC to select the optimal number of subtypes. **c** The probability of subtype assignment across SuStaIn subtypes. ****P* < 0.001. **d** The probability of subtype assignment at each SuStaIn stage of the two molecular subtypes. SuStaIn = Subtype and Stage Inference; CVIC = cross-validation information criterion; S0 = Normal-appearing group; S1 = Immune/Apoptosis/Proteostasis subtype; S2 = Synapse/RNA-Metabolism subtype.

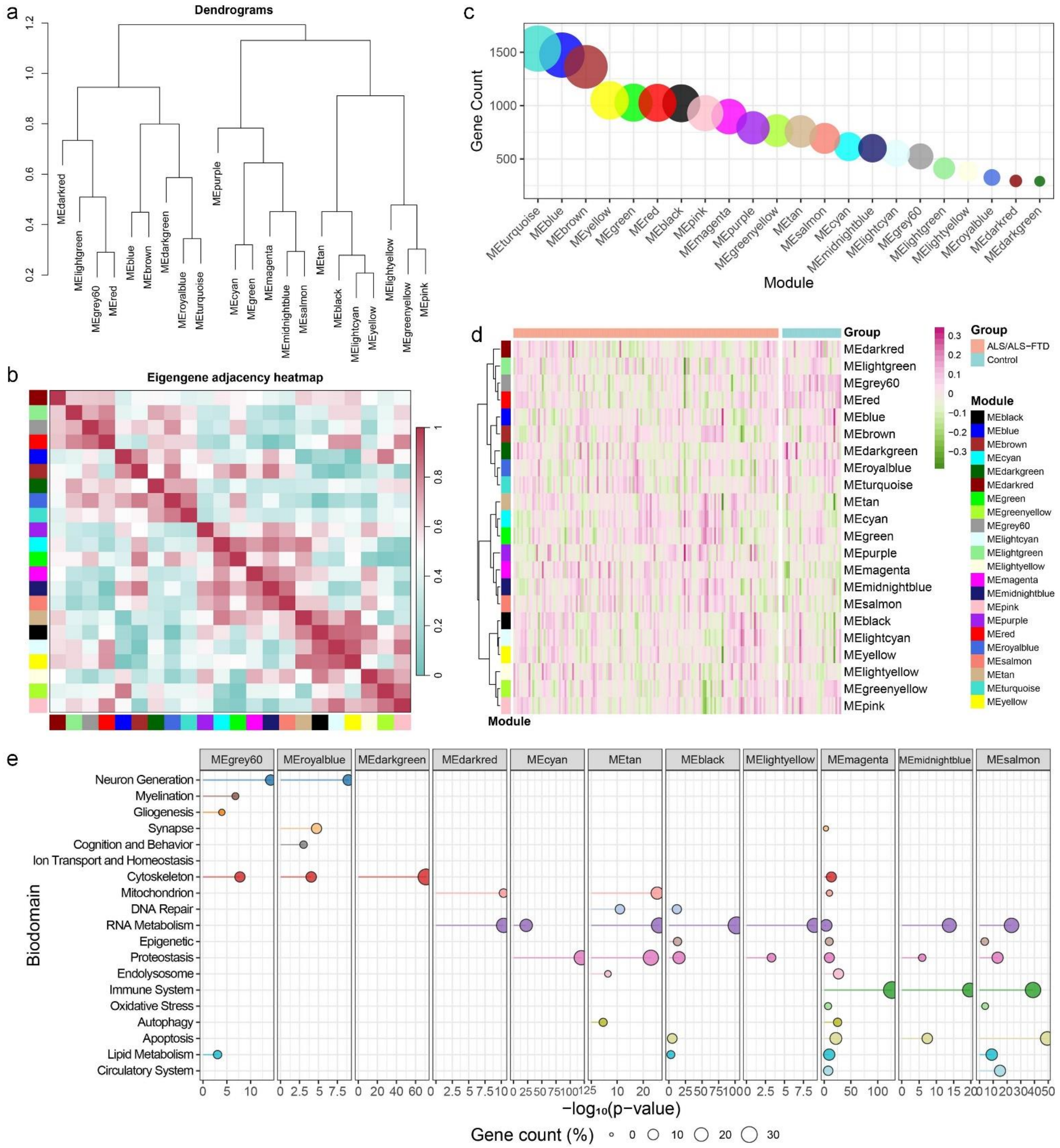

**Supplementary Fig. 3 Co-expression module construction in lumbar spinal cord samples. a** Hierarchical clustering dendrogram illustrating the relationships between different modules. **b** This heatmap visualizes the relationships between eigengenes of identified co-expression modules. Each cell in the heatmap represents the correlation between the eigengenes of two different modules, with color intensity indicating the strength of the relationship. **c** The size of each modules. **d** The module eigengenes for each module within individual samples. **e** Biological annotation of each module. Each dot represents the genes associated with a specific biodomain. Color of the dots represent different biodomains. The x-axis represents the significance level of the top GO term linked with each biodomain, with more significant terms positioned further to the right. The size of each dot corresponds to the proportion of genes related to that biodomain within each module.

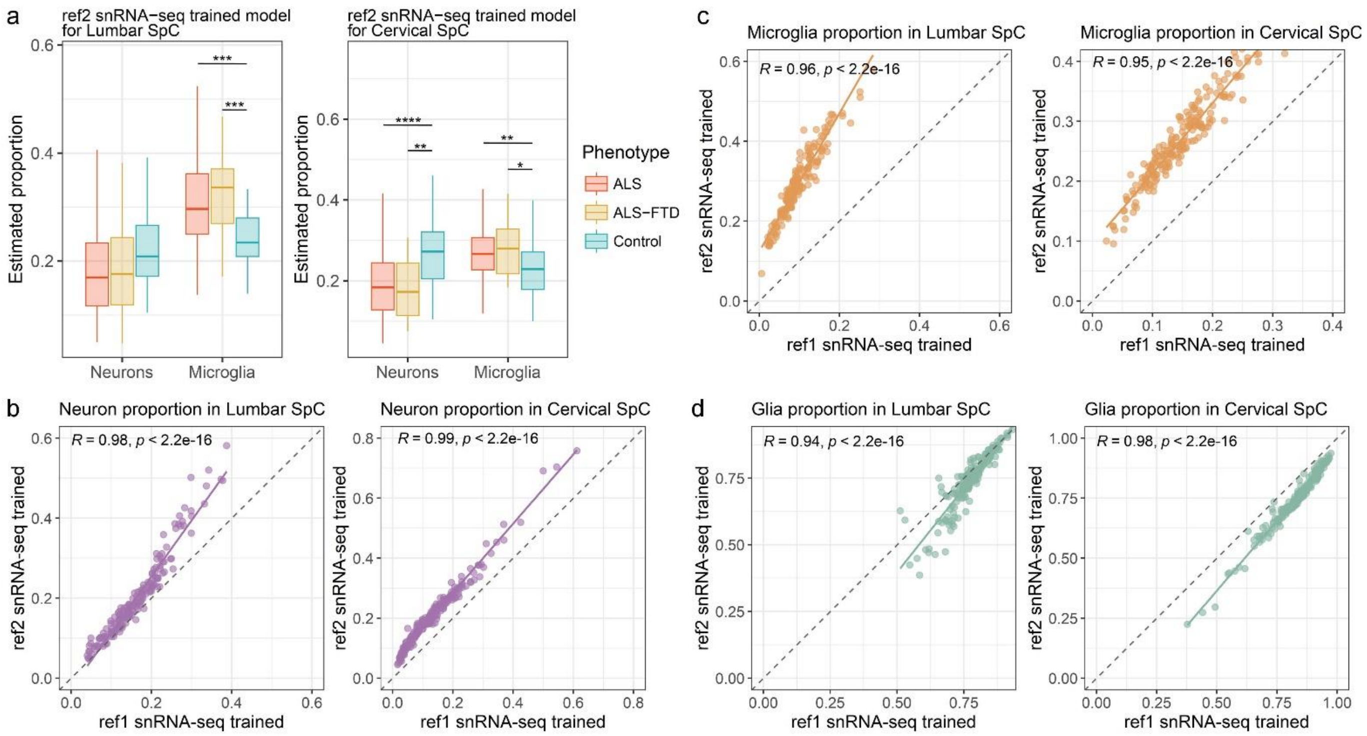


**Supplementary Fig. 4 Distributions of cell-type proportions. a** Cell-type proportions in lumbar and cervical across different clinical phenotypes. This Scaden model was trained by using the additional snRNA-seq as reference. *$P < 0.05$, **$P < 0.01$, ***$P < 0.001$, ****$P < 0.0001$. **b-d** Correlations between estimated cell-type proportions generated using two snRNA-seq references.

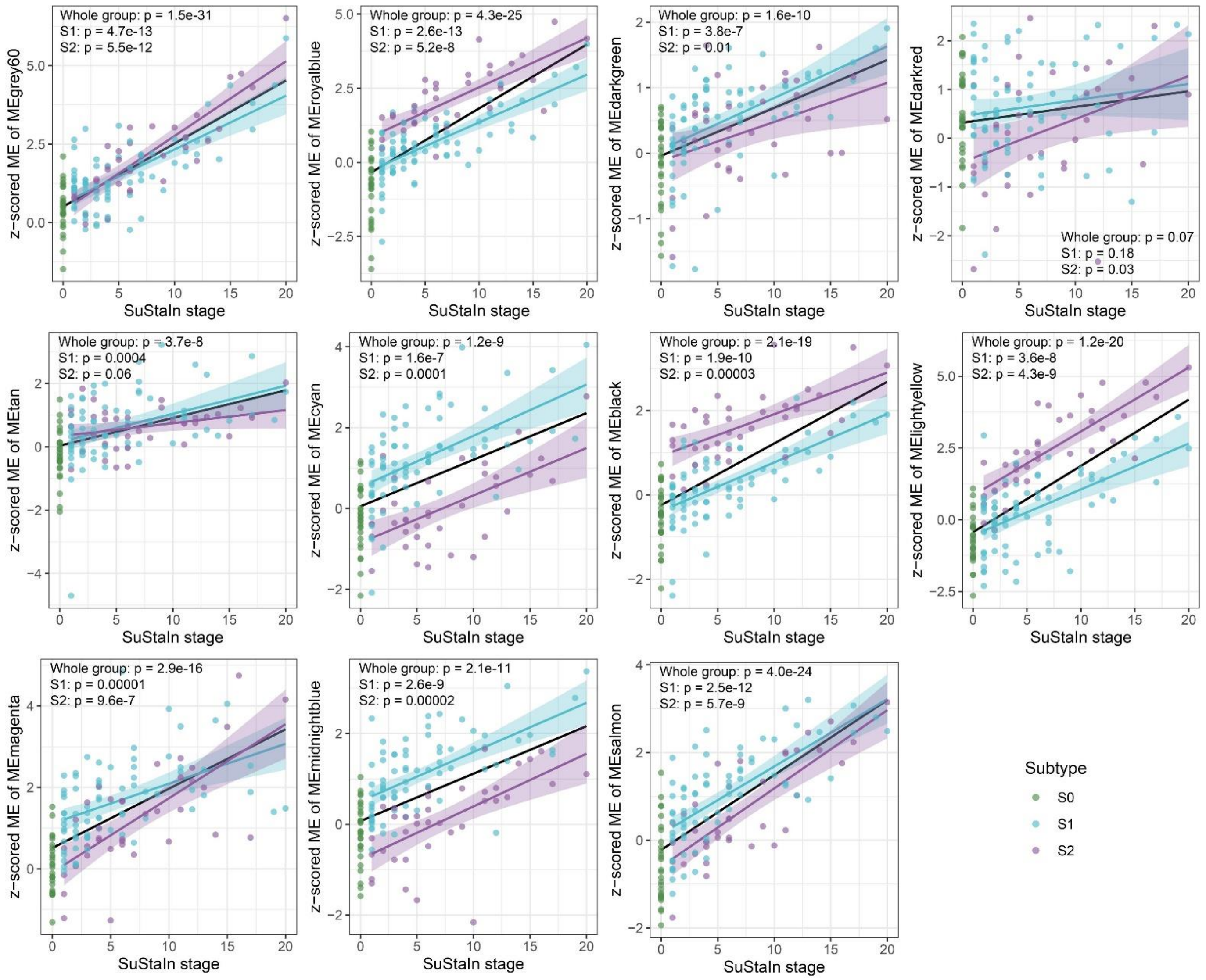


**Supplementary Fig. 5 SuStaIn stage correlated with z-scored MEs of core modules.** SuStaIn = Subtype and Stage Inference; S0 = Normal-appearing group; S1 = Immune/Apoptosis/Proteostasis subtype; S2 = Synapse/RNA-Metabolism subtype; ME = module eigengene.

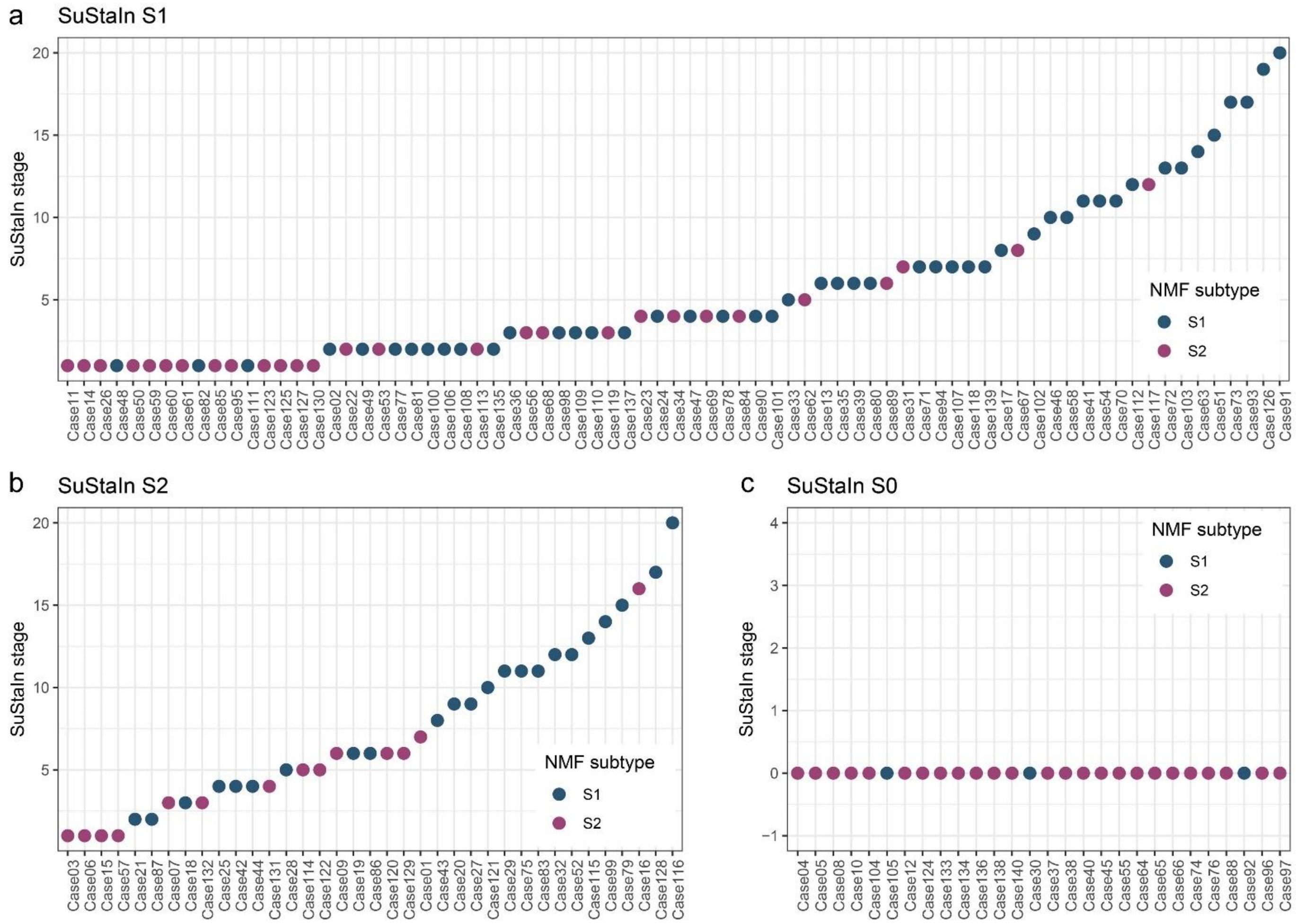


**Supplementary Fig. 6 SuStaIn subtypes at different stages mapping to NMF subtypes.** SuStaIn = Subtype and Stage Inference; SuStaIn S0 = Normal-appearing group; SuStaIn S1 = Immune/Apoptosis/Proteostasis subtype; SuStaIn S2 = Synapse/RNA-Metabolism subtype; NMF = non-negative matrix factorization; NMF S1 = Immune and RNA/Protein metabolism subtype; NMF S2 = Synapse subtype.

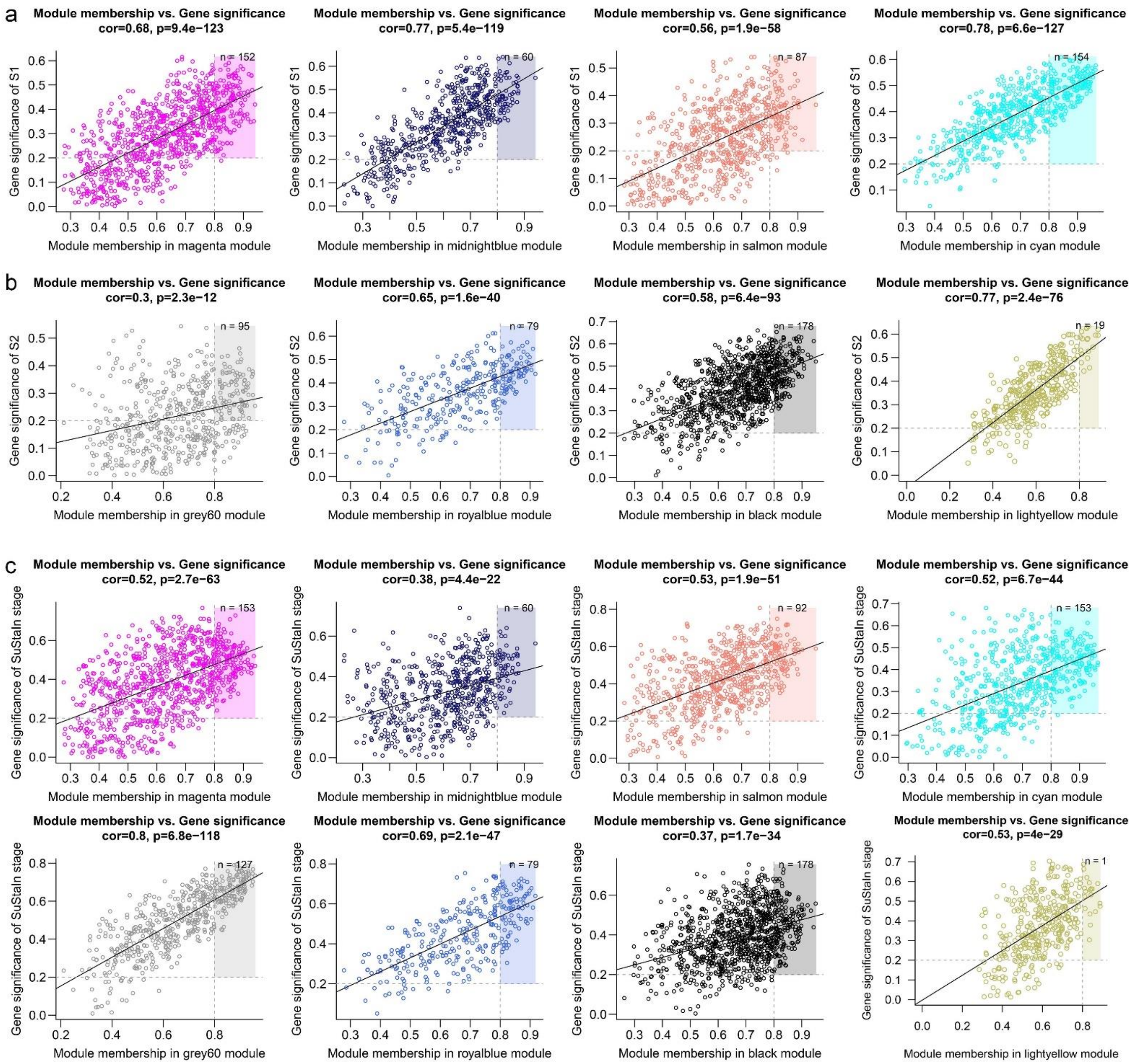


**Supplementary Fig. 7 Hub genes within core modules associated with subtyping and staging. a** The core modules associated with the Immune/Apoptosis/Proteostasis subtype (S1). **b** The core modules associated with the Synapse/RNA-Metabolism subtype (S2). **c** The core modules associated with SuStaIn stage. Scatter plots of GS score and MM for genes in each module. Genes with GS > 0.2 and MM > 0.8 were highlighted. GS = gene significance; MM = module membership.

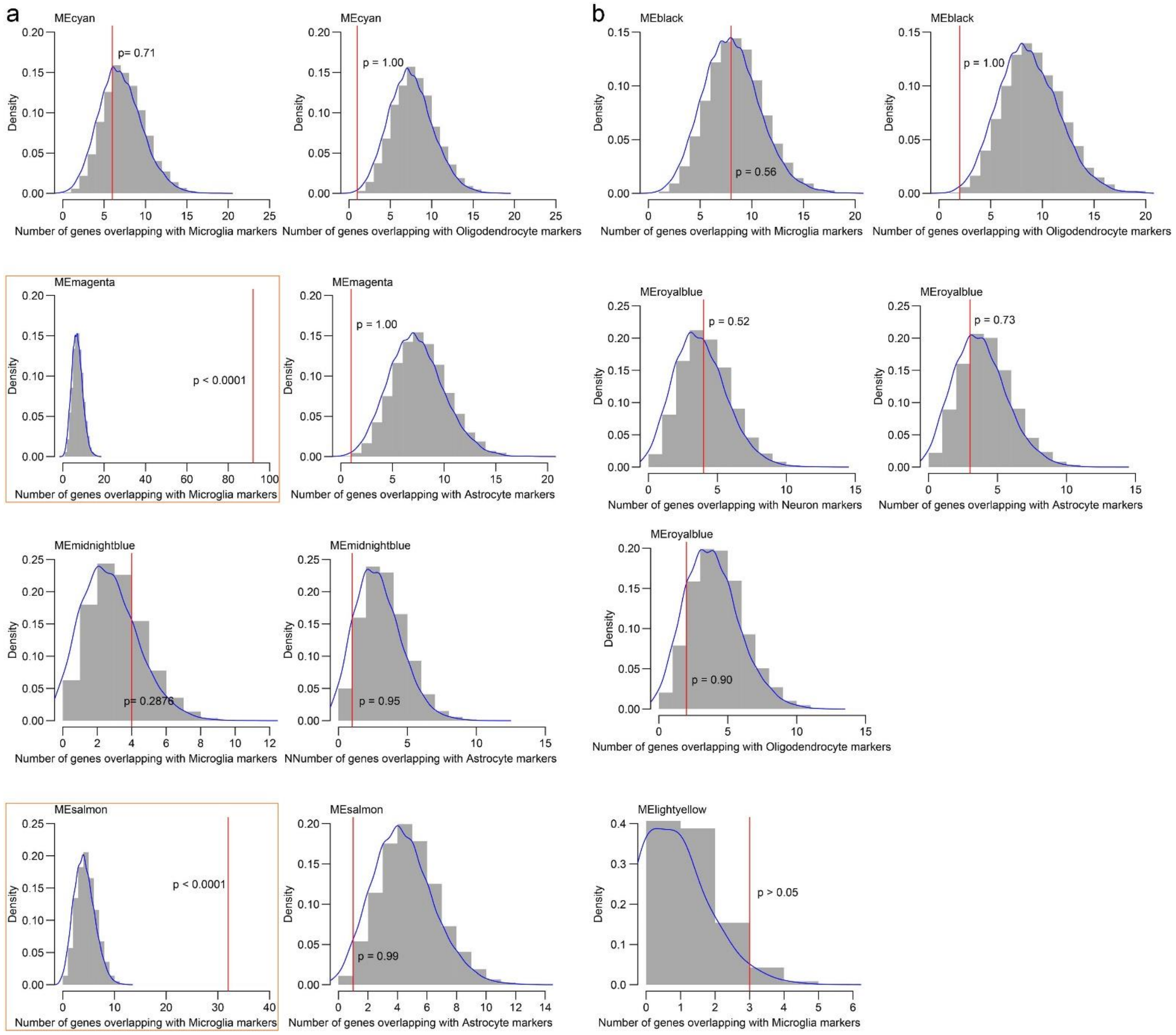


**Supplementary Fig. 8 The overlap between hub genes within each module and cell-type marker genes.** **a** The core modules associated with the Immune/Apoptosis/Proteostasis subtype (S1). **b** The core modules associated with the Synapse/RNA-Metabolism subtype (S2). Histograms display the null distribution, depicting the number of overlapped genes. The red line represents the observed number of overlapped genes. Results highlighted with an orange border denote those that reached statistical significance.

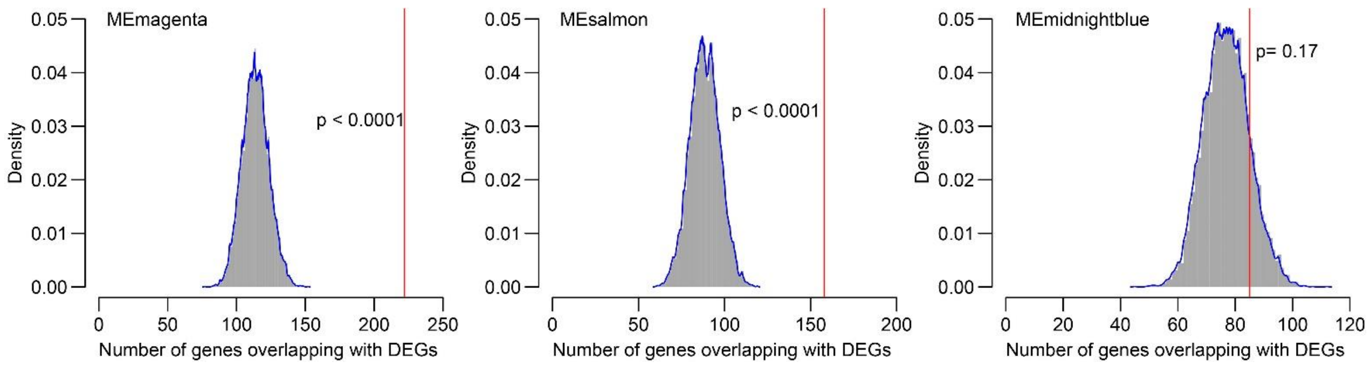


**Supplementary Fig. 9 The overlap between all genes within each module and treatment-responsive genes.** Histograms display the null distribution, depicting the number of overlapped genes. The red line represents the observed number of overlapped genes.
